# Wild spirits: Elevated hypomanic tendencies are associated with entrepreneurship and entrepreneurial success

**DOI:** 10.1101/2020.05.17.20104836

**Authors:** Helen Pushkarskaya, Christopher Pittenger, Marco Kleine, Godfrey Pearlson, David R. Just

## Abstract

Entrepreneurs take extreme risks and make rash decisions but are responsible for innovation, technological change, and economic growth. Attempts to characterize the entrepreneurial personality have had limited success. We hypothesized that elevated hypomanic tendencies (HPM), which have been linked to increased risk for psychopathology, are also associated with entrepreneurship. We tested the association of HPM with entrepreneurial activities cross-sectionally in three different populations (total N = 2,200, including 899 entrepreneurs), using three different measures of hypomanic tendencies. We report a robust positive association of HPM with several measures of entrepreneurial intent, behavior, and success. This may reflect a particular ecological fit between individual characteristics associated with HPM and unique demands of entrepreneurial lifestyle, but highlights the psychological vulnerability of entrepreneurs, especially under conditions of stress.

**One Sentence Summary:** A positive relationship between hypomanic tendencies and entrepreneurship is revealed in three large independent datasets.

## Main Text

Joseph Schumpeter, one of the most influential economists of the 20^th^ century, described entrepreneurs as “wild spirits” who take extreme risks and make rash decisions but are responsible for innovation and technological change, and thus for economic growth (*1*). Much subsequent literature has attempted to characterize the entrepreneurial personality and to identify determinants of entrepreneurial drive (*2*). Entrepreneurship is not associated with the dimensions of personality in the prominent ‘Big 5’ personality model (*2*). However, many of the documented characteristics of entrepreneurs (*3-6*) parallel the older construct of hypomanic personality (*7-13*): impulsivity (*14, 15*), altered perception of risk (*16*) and increased risk taking (*17, 18*), decreased tendency to think counterfactually (*19*), ambition and overconfidence (*20*), perseverance (*21*), positive affectivity (*22*), goal orientation (*23, 24*), little need for sleep (*25, 26*), and reduced regard for social norms (*27, 28*). Mania and hypomania have long been associated with creativity and with increased goal-directed activity (*7*, *10, 29-37*). There have been many case studies suggesting a relationship between hypomanic tendencies and entrepreneurship (*7-9, 32*); but the empirical literature is limited and mixed. Some small studies have supported an association between hypomania and entrepreneurship (*32*); others have found no such relationship (*33*). These small studies have used a range of measures of hypomania and of entrepreneurial behavior. We hypothesized that strong and persistent hypomanic traits (HPM) would be associated with entrepreneurship. We tested the association of HPM with entrepreneurial activities cross-sectionally in three different populations (total N = 2,200, including 899 individuals with a history of business creation), using three different measures of hypomanic traits (detailed below). We report a robust positive association of HPM with several different measures of entrepreneurial intent, behavior, and success. Our results thus provide empirical support for the “wild spirits” hypothesis of entrepreneurship and provide new insight into the entrepreneurial drive.

“Good science has to begin with good definitions” (*34*). Some have defined entrepreneurship as the creation of any new business (*35*), while others restrict it to the creation of innovative businesses, excluding the creation of ventures that are not innovative (like opening a franchise) (*36*). In either case, the success rate of new ventures is very low, across a wide array of success measures (*37-42*). Some researchers also include innovation within an existing and thriving corporation (*42, 43*) or in the sphere of social activism (*44*) under the umbrella of entrepreneurship. We examine several of these different notions of entrepreneurship but focus primarily on the creation of new ventures (irrespective of the level of innovation).

Hypomania has likewise been conceptualized and measured in a variety of ways (*45-49*). It is a risk factor for the development of bipolar disorder (~20%, versus ~1.5% in general population) (*50, 51*), but most individuals with HPM do not go on to develop major psychopathology (*52, 53*). Some studies have documented the mental health sequelae of the stress of venture creation (*54-56*). We posit that psychiatric difficulties in this population may derive from an interaction of this stress with an underlying temperamental vulnerability (i.e., HPM). However, we emphasize that the very factors associated with this vulnerability may simultaneously serve an important adaptive purpose, driving the creation of technological change and economic growth (*57*).

Whether mania exists on a continuum of severity or not, with hypomanic personality representing a mild form, depends in part on the type and level of analysis. This is one aspect of the broader question of whether continuous personality traits (e.g. hypomanic tendencies) can be truly separated from discrete pathologic entities (e.g. psychiatric illnesses such as bipolar disorder). Both biological measures (e.g. polygenic risk scores for bipolar disorder (*58*)) and non-biological measures (e.g. family pedigree analysis of bipolar family members (*59*)) may help resolve the question of whether hypomania and bipolar disorder lie on a continuum. We did not here attempt to resolve this important question.

We tested the association of HPM with entrepreneurship in three independent studies: a sample of 1387 residents of Kentucky (a primarily rural state in the U.S.), which was oversampled to include 45% (619) who reported having created a business (Study 1; Supplementary text); a sample of 417 general population respondents recruited in Connecticut, 72 of whom reported a history of creating a business (Study 2; Supplementary text); and an international group of 396 individuals from executive MBA programs, 208 of whom described themselves as having created a business (Study 3; Supplementary text). In all three samples, using different instruments to measure HPM, individuals with strong hypomanic propensities were overrepresented among individuals with a history of business creation, especially among individuals with a history of creating multiple businesses (i.e. serial entrepreneurs; Figure 1). This robustly supports the hypothesized positive association between HPM and entrepreneurship.

**Fig. 1.**
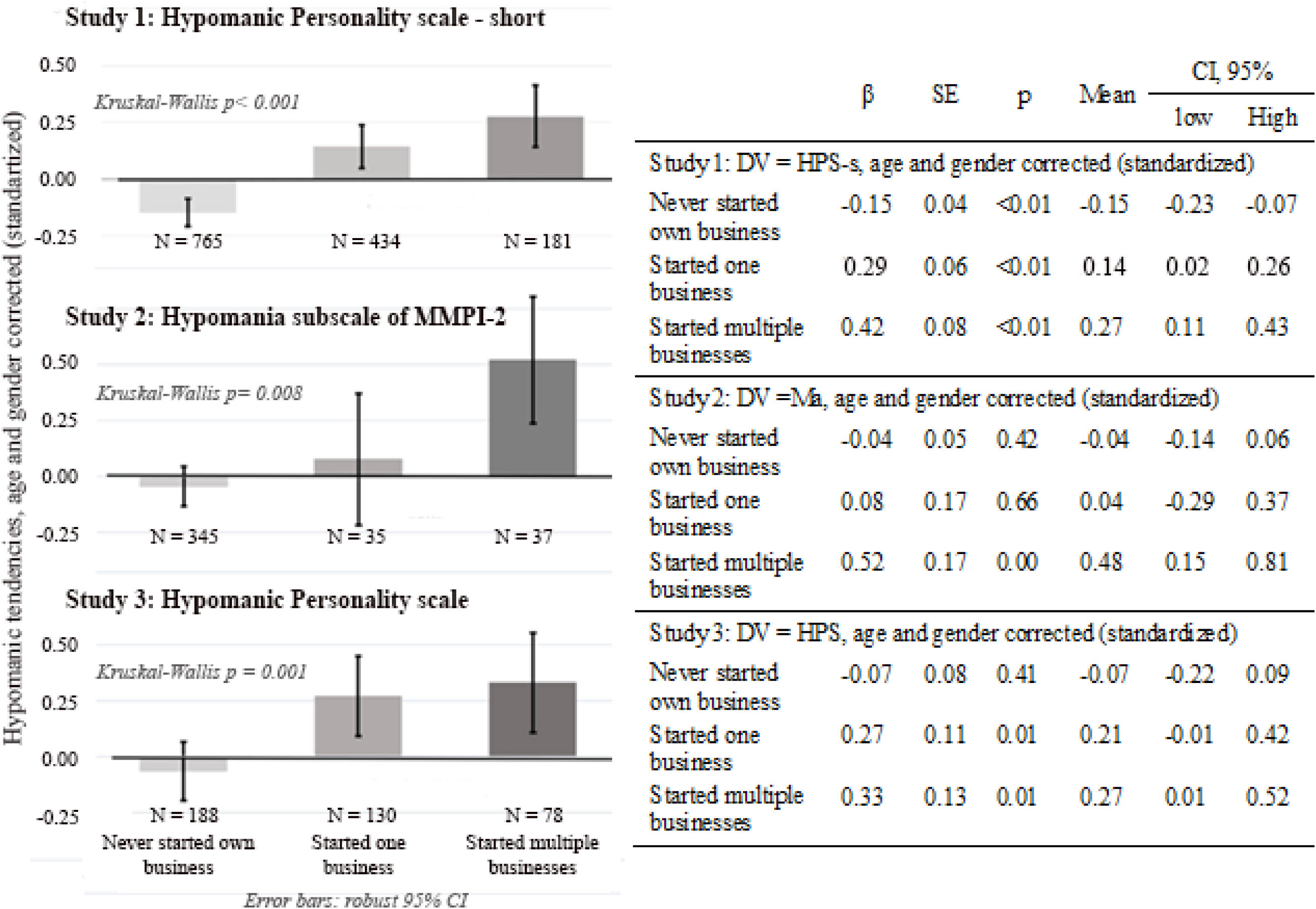
Relationship between history of entrepreneurial activities and hypomanic personality, across three studies. We collected data in three different samples, totaling 2200 individuals, and assessed hypomanic tendencies using three different instruments. In all three cases there was a monotonic relationship between history of business creation (never, once, more than once) and hypomanic tendencies. Parameter estimates (βs) and standard errors (SE) are from a regression with a heteroskedasticity-consistent variance covariance matrix (robust linear regression).

The Hypomanic Personality Scale (HPS) of Eckblad and Chapman (*46*) is the most widely used instrument to measure hypomanic tendencies in non-clinical samples, but it is inconveniently long for large-scale surveys. In Study 1 (N = 1387), we measured HPM using a shortened version of the HPS (HPS-short, or HPS-s), which we validated against the full HPS in an independent group of subjects (Supplementary text, Figure S1). Entrepreneurial attitudes were evaluated using two validated scales: the Business Risk-Taking Scale (BRT) (*60*) and a shortened version of the Entrepreneurial Intentions Scale (EI) (*61*). Details of these scales, including the construction and validation of the shortened EI (EI-r), are given in Supplementary text, as are demographic characteristics of the sample and summary statistics of measures of HPM and entrepreneurship.

As shown in Figure 1A, there was a positive and monotonic relationship between HPS-S and history of business creation: serial entrepreneurs on average scored higher than one-time entrepreneurs, who in turn on average scored higher than individuals that never created a business. Similar significant effects were seen when we used a dichotomous measure of business entrepreneurship (i.e. “have you ever started a business [yes/no]”; Figure S2).

In this large sample we were able to examine association of HPM with <u>current</u> entrepreneurial activity and intentions, in addition to history. Hypomanic propensities were elevated in those with any past or present business creation, relative to those who had never started a business, but were most markedly elevated in those who were actively in the process of starting a business (nascent entrepreneurs, NE; Figure 2A; see Supplementary text) (*62-64*). This could be because elevated HPM represents a stable trait, and individuals with high HPM self-select into the current entrepreneur group (*65*). Alternatively, HPM may represent a combination of stable trait and more transient state effects (*66*); individuals may be drawn into venture creation during periods when their HPM is high, or elevated HPM may be triggered, in susceptible individuals, by the stress of venture creation. Our cross-sectional design does not allow us to distinguish between these possibilities.

**Fig. 2.**
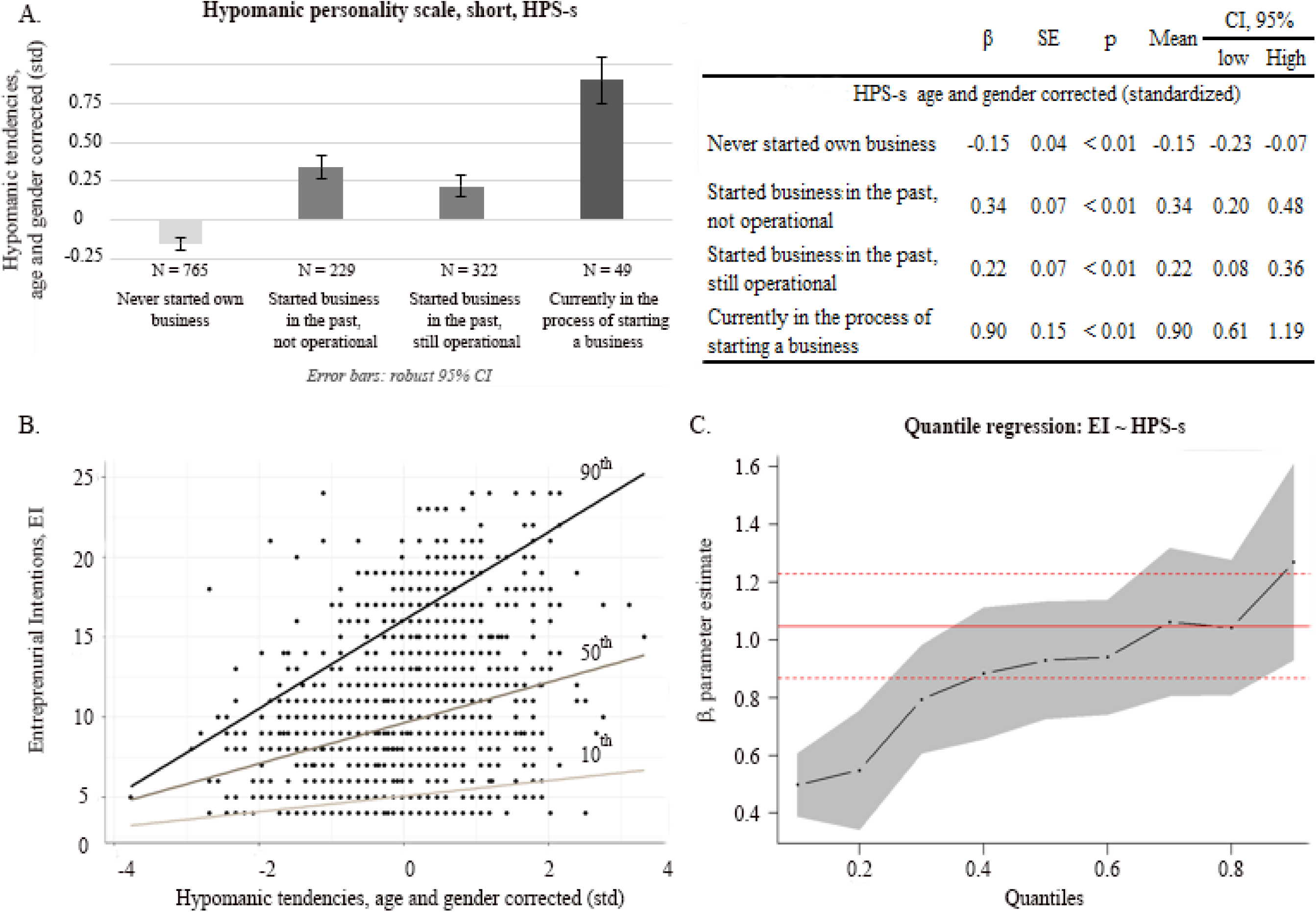
Association of hypomania with entrepreneurial intentions and current entrepreneurial activity in Study 1. A. HPM was elevated in all individuals who had a history of business creation, relative to those who did not, but was most markedly elevated in individuals currently in the process of starting a new business (nascent entrepreneurs). Parameter estimates (βs) and standard errors (SE) are from a regression with a heteroskedasticity-consistent variance covariance matrix (robust linear regression). B. There was a positive relationship between hypomania and current entrepreneurial intentions (EI). Regression lines are shown for the 10%, 50%, and 90% quantiles. C. Regression coefficients (in black) and confidence intervals (in gray) across all quantiles. In red are a coefficient (solid line) and confidence intervals (dotted lines) from the ordinary linear regression.

Similarly, there was a positive association between hypomanic propensities and current entrepreneurial <u>intentions</u> (irrespective of actual activity), measured using the EI-r (Figure 2B). Since the variance in EI-r increased dramatically at higher levels of HPS-s (Breusch–Pagan test p < 10^-10^), we analyzed these data using quantile regression (*67*), which divides EI-r into nine quantiles at each level of HPS-s, and then examines the relationship between HPS-s and EI-r in each of these subsets of the data.^1^ As shown in Figure 2C, the relationship between HPS-s and EIr was significant and positive across all quantiles, but stronger in the higher quantiles (see also Table S9). That is, not all individuals with HPM in our sample formed entrepreneurial intentions, but almost no individuals without HPM did so.

In aggregate, these several analyses in the initial cohort of over 1,000 show a robust positive association of HPM with entrepreneurial behavior and intention.

In Study 2, we examined the specificity of the association of HPM with venture creation, relative to different personality constructs and different conceptions of entrepreneurship. Hypomanic propensities do not map cleanly onto the most widely used dimensional model of personality, the Big Five model, and no associations have been found between entrepreneurship and the dimensions of that model (*2*). Hypomania is specifically measured by an older, but extremely well validated and widely used, instrument for characterizing personality variation and pathology along nine dimensions: the Minnesota Multiphasic Personality Inventory (MMPI) (*49, 68*). We used a more recent version of MMPI, MMPI-2 (*49*), and several measures of entrepreneurship (as defined by the entrepreneurial outcomes inventory, EO (*69*)) to access 417 general population subjects, 72 of whom had a history of creating a business (see Supplementary text for details of these measures and descriptive statistics of the sample). As in Study 1, hypomanic tendencies (the MMPI-2 Ma subscale) were positively associated with history of business creation, measured either across three levels (Figure 1B) or as a dichotomous variable (Table 1). The other eight MMPI-2 dimensions showed no such relationship (Table 1; Supplementary text, Table S14). Thus, the positive association of venture creation with hypomanic propensities is specific, at least amongst the nine dimensions measured by the MMPI-2.

**Table 1.**

|  | **Business** |  |  | **Corporate** |  | **Invention** |  |
| --- | --- | --- | --- | --- | --- | --- | --- |
|  | **Entrepreneurship** | **Student Entrepreneurship** |  | **Entrepreneurship** |  | **Entrepreneurship** |  |
|  | **Mann-Whitney U test,** |  |  |  |  |  |  |
|  | **p** | **Spearman's $\rho$** | **p** | **Spearman's $\rho$** | **p** | **Spearman's $\rho$** | **p** |
| Hs | 0.310 | -0.03 | 0.534 | 0.02 | 0.680 | 0.02 | 0.703 |
| D | 0.180 | -0.14 | 0.006 | -0.19* | <0.001 | -0.09 | 0.077 |
| Hy | 0.800 | < 0.01 | 0.990 | 0.01 | 0.830 | 0.03 | 0.589 |
| Pd | 0.960 | -0.13 | 0.011 | -0.05 | 0.355 | -0.06 | 0.210 |
| Pa | 0.750 | -0.03 | 0.534 | -0.01 | 0.847 | 0.00 | 0.937 |
| Pt | 0.920 | -0.09 | 0.057 | -0.10 | 0.036 | -0.07 | 0.178 |
| Sc | 0.200 | -0.05 | 0.352 | -0.03 | 0.555 | -0.05 | 0.291 |
| Ma | 0.006* | 0.15* | 0.002 | 0.18* | <0.001 | 0.05 | 0.267 |
| Si | 0.150 | -0.18* | <0.001 | -0.21* | <0.001 | -0.12 | 0.012 |

Relationship between MMPI dimensions and several types of entrepreneurship in Study 2. \* - significant at a level corrected for multiple comparisons: for Mann-Whitney U tests $p \leq (0.05/9) = 0.006$ ; for Spearman's correlations $p \leq (0.05/27) = 0.002$ .

Hitherto we have defined entrepreneurship as a history of business creation. However, entrepreneurship has been defined and studied in other ways. We examined the association of other forms of entrepreneurship with MMPI-2 personality dimensions. Significant relationships were seen between the MMPI-2 Ma subscale and both student and corporate entrepreneurship (Table 1). Negative associations emerged with MMPI-2 D and Si subscales (measuring depression and social isolation tendencies, respectively) and student and corporate entrepreneurship – that is, these dimensions work against these forms of entrepreneurship. Overall, across several different definitions of entrepreneurship, MMPI-2 Ma has the most consistent association, and the only positive one, further supporting the robustness and specificity of the relationship. There were no significant associations between any MMPI-2 dimensions and innovation entrepreneurship.

In Study 3 we tested whether the association between hypomanic tendencies and entrepreneurship would hold in individuals who self-selected to receive business education, and how it associates with entrepreneurial success. We used on-line data collection to characterize an international group of 396 individuals enrolled in executive MBA programs, currently or in the past; 208 described themselves as having created a business (Supplementary text; Table S15). Hypomanic tendencies in this sample were characterized using the full HPS. We again replicated the core finding seen in the first two studies: that a history of having started a business associated positively with HPM (Figure 1C).

Collection of the full HPS in this sample allowed us to examine three well-validated subscales that measure different aspects of HPM (70): social vitality (SV), mood volatility (MV), and excitement (Ex; see Supplementary text). These subscales are highly intercorrelated in general, and in our sample (Supplementary text). History of business creation was positively associated with SV (mean_one_ – mean_none_ = 0.23, mean_several_ – mean_one_ = 0.24; Kruskal-Wallis p = 0.001) and, more weakly, with Ex (mean_one_ – mean_none_ ^=^ 0.23, mean_several_ – mean_one_ ^=^ 0.01; Kruskal-Wallis p = 0.014); there was no consistent monotonic association with MV (mean_one_ – mean_none_ = 0.23, mean_several_ – mean_one_ = −0.12; Kruskal-Wallis p = 0.060; Table 2). This suggests that the association between hypomanic propensities and entrepreneurship is driven primarily by social vitality. Relationships of HPS subscales with the other forms of entrepreneurship measured by the EO were weaker (Table S17).

**Table 2.**

| | $\beta$ | SE | p | Mean | CI, 95% | |
| --- | --- | --- | --- | --- | --- | --- |
|  |  |  |  |  | low | high |
| *DV = SV, age and gender corrected (standardized)* |  |  |  |  |  |  |
| Never started own business | -0.13 | 0.08 | 0.120 | -0.13 | -0.29 | 0.03 |
| Started one business | 0.23 | 0.11 | 0.040 | 0.10 | -0.12 | 0.32 |
| Started multiple businesses | 0.47 | 0.13 | <0.01 | 0.34 | 0.09 | 0.59 |
| *DV = MV, age and gender corrected (standardized)* |  |  |  |  |  |  |
| Never started own business | 0.01 | 0.05 | 0.778 | 0.01 | -0.09 | 0.11 |
| Started one business | 0.23 | 0.16 | 0.522 | 0.24 | -0.07 | 0.55 |
| Started multiple businesses | 0.11 | 0.18 | 0.035 | 0.12 | -0.23 | 0.47 |
| *DV = Ex, age and gender corrected (standardized)* |  |  |  |  |  |  |
| Never started own business | -0.04 | 0.08 | 0.590 | -0.04 | -0.20 | 0.12 |
| Started one business | 0.23 | 0.11 | 0.040 | 0.19 | -0.03 | 0.41 |
| Started multiple businesses | 0.24 | 0.13 | 0.060 | 0.20 | -0.05 | 0.45 |

Association of history of entrepreneurship with HPS subscales. SV – Social Vitality; MV – Mood Volatility; Ex – Excitement.

A subset of this third sample (N = 109) provided data on past entrepreneurial success, measured as peak annual revenue of the most successful business created. We examined the relationship of business success with the three HPS subscales (Supplementary text; Table S17). Since business success is highly skewed – high levels of success are rare (*36-39*) – analyzing this relationship again required the use of quantile regression (67), including as predictors the three HPS subscales and their squares (to allow for quadratic relationships). Only SV predicted entrepreneurial success, and only in the top quantile (Figure 3; Table S18): that is, most entrepreneurial enterprises fail, but amongst those that succeed, social vitality of the entrepreneur is a significant predictor of the degree of success, as measured by peak income.

**Fig. 3.**
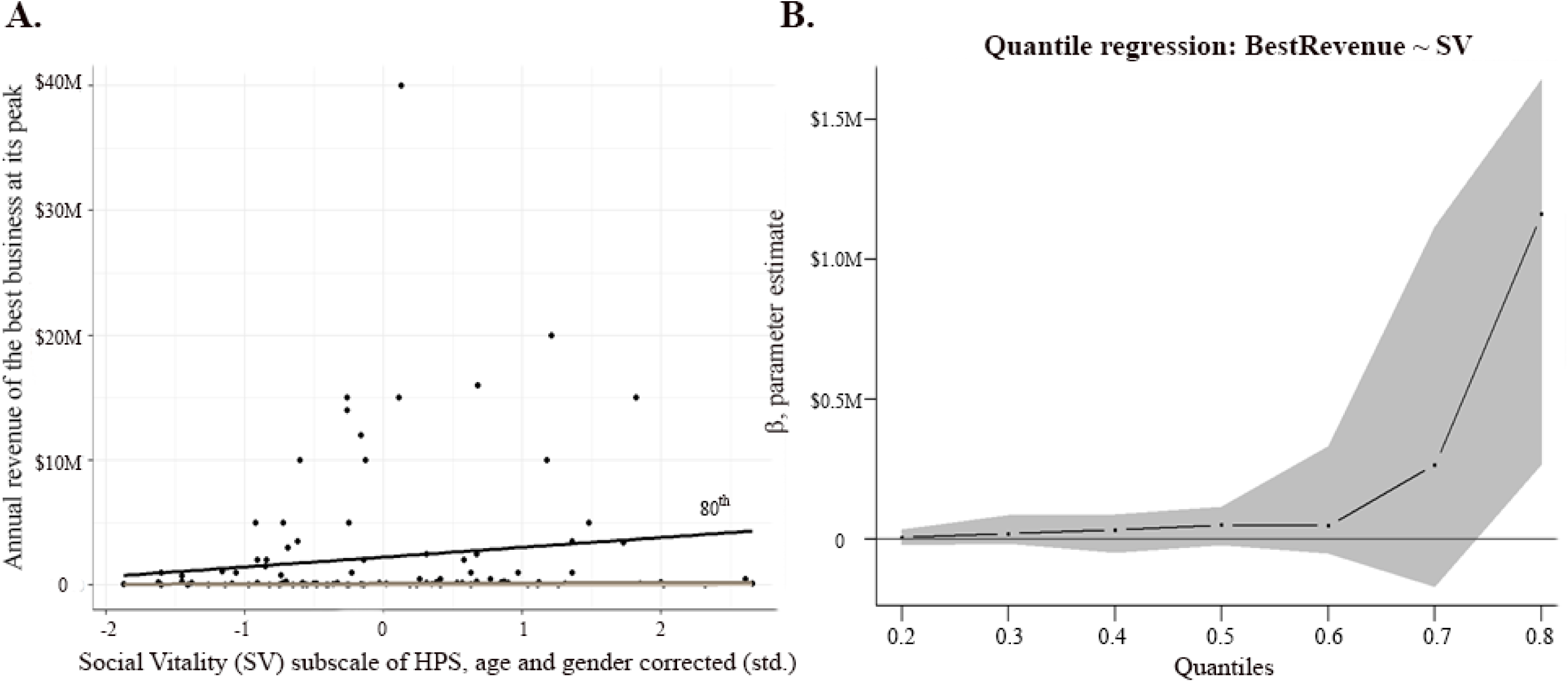
Quantile regression of the relationship between social vitality (SV) and entrepreneurial success in Study 3. Of the HPS subscales, only SV was associated with entrepreneurial success, defined as peak revenue of businesses started (Table S6.2). Most business ventures fail, and thus at lower quantiles there was little variation in peak revenue and no correlation with SV. In the top quantiles, however, a positive relationship emerged.

A minority of economic actors function as entrepreneurs, and yet the actions of this subgroup are critical to innovation and economic growth (*1*). It has been challenging to identify personality traits of individuals who will self-select to function as entrepreneurs in the economy, and who will be successful. We provide convergent data across three large samples, supporting a positive association between hypomanic tendencies and entrepreneurship. Notably, HPM is particularly high among serial entrepreneurs – individuals who select venture creation as a lifestyle rather than a onetime activity – and among nascent entrepreneurs, as they navigate a particularly stressful period.

Extreme personality traits are often thought to be associated with psychopathology and impaired function. HPM is no exception; elevated HPM is a predictor of the development of bipolar disorder (*50, 51*). However, it has been speculated that HPM (and even bipolar disorder) may have benefits as well; for example, case reports and some studies associate HPM with creativity (*30, 31*). We provide evidence that elevated HPM also contributes positively to entrepreneurship. This may reflect a particular ecological fit between individual characteristics associated with HPM and unique demands and opportunities of entrepreneurial lifestyle (*71*). This association is specific; none of the other measured personality dimensions correlate with business entrepreneurship. Social vitality (SV) appears to be the strongest predictor of both entrepreneurial activity and success. However, SV strongly correlates with other dimensions of HPM, mood volatility and excitement; this combination of intercorrelated traits may constitute the “wild spirit” of entrepreneurs.

Improved understanding of the relationship between personality variation and entrepreneurship may help policy makers, educational institutions, and support programs to identify, cultivate, and support potential and active entrepreneurs. The identification of a psychological trait (which has been suggested to be heritable (*72*)) that is associated both with entrepreneurship and with psychopathology highlights the psychological vulnerability of entrepreneurs and their family members (*73*), especially under conditions of stress. However, it is important to emphasize that most individuals with HPM (~80%) do not go on to develop major psychopathology (*52, 53*). Identifying resilience factors in this high-risk population is a critical topic for future research. Hypomania may be a double-edged sword. Appreciation of this fact may inform strategies to support entrepreneurs as they provide a critical engine for the economy.

## Data Availability

All de-identified data and R scripts are available at https://github.com/helenpush/HPM.git .

## Acknowledgments

The authors extend sincere thanks to Dr. Hal Arkes of Ohio State University for constructive and thoughtful comments on the manuscript; to Drs. Nicole Breazeale, Alison Davis, Ellen Usher, and Ronald Hustedde of University of Kentucky who were essential for the collection of data for Study 1; Jessica Halten of Yale School of Management who was essential for collection of data for Study 2; critical graduate student support for Study 1 was provided by Cami Bush at Western Kentucky University (WKU), Shaheer Burney of CEDIK, Spencer Walters and Stephen Gibbons of WKU. The authors express gratitude to The Community and Economic Development Initiative of Kentucky (CEDIK) for supporting Study 1; Behavioral Laboratory of Yale School of Management for supporting Study 2; LMU Entrepreneurship Center for distributing the Study 3 survey via their network.

## Author contributions

Helen Pushkarskaya: methodology, software, validation, formal analysis, investigation, resources, data curation, writing – review and editing, visualization, supervision, project administration, funding acquisition; Christopher Pittenger: methodology, validation, writing – original draft preparation, writing – review and editing, supervision, Marco Kleine: software, validation, investigation, resources, data curation, writing – review and editing, project administration; Godfrey Pearlson: validation, writing –review and editing, supervision; David R. Just – conceptualization, methodology, validation, formal analysis, resources, writing – review and editing, supervision;

## Data and materials availability

All data is available in the main text or the supplementary materials.

## Supplementary Materials

Materials and Methods

Supplementary text

Figures S1-S3

Tables S1-S18

External Databases S1-S9

References (74-106)

## Other Supplementary Materials for this manuscript include the following

Data S1. Study1.csv - Input file for non-parametric analysis in R, Study 1.

Data S2. Study1.sav - Full dataset, Study 1.

Data S3 Study2.csv - Input file for non-parametric analysis in R, Study 2.

Data S4. Study2.sav - Full dataset, Study 2.

Data S5. Study3.csv -Input file for non-parametric analysis in R, Study 3.

Data S5a. Study3R.csv - Input file for non-parametric analysis in R, Study 3 - subsample for analysis of business success.

Data S6. Study3.csv - Full dataset, Study 3.

Data S7. Study_1_R_output.html - Code and output for nonparametric analyses in R, Study 1.

Data S8. Study_2_R_output.html - Code and output for nonparametric analyses in R, Study 2.

Data S9. Study_3_R_output.html - Code and output for nonparametric analyses in R, Study 3.

## Materials and Methods

To empirically examine relations between entrepreneurial behavior and individual hypomanic tendencies, we conducted three studies (as detailed below). We collected three large datasets (N = 1387, 417, 396), from geographically and socioeconomically distinct samples. We used three different measures to quantify hypomanic propensities, and six different measures to quantify entrepreneurial behavior.

### Approvals of institutional review boards

The respective local ethics committees (Institutional Review Board at University of Kentucky, and Yale University Institutional Review Board) approved each study, which were conducted in accordance with the Declaration of Helsinki. The anonymous mailed-in survey (Study 1) was judged to be exempt from informed consent. For computer-based data collection (Studies 2 and 3), an approved informed consent form was presented on the first page. Only after participants indicated agreement to participate in the study on this form did they proceed to the surveys.

### Instruments

HPM was measured using three different instruments. In Study 3 we used the well-validated Hypomanic Personality Scale (HPS) of Eckblad and Chapman (46); further details about this scale are given in the Supplementary Text. For Study 1 we constructed and validated a shortened version of this scale, HPS-s, which we found to correlate well with the HPS; further details of this scale, including the procedure used to generate and validate it and the complete item list, are given in the Supplementary Text. In Study 2 we used the Minnesota Multiphasic Personality Inventory-2 (MMPI-2 (49)) to more comprehensively assess personality dimensions; the hypomania scale (Ma (74)) was used as the measure of hypomania in the primary analyses, while other scales were used to assess the specificity of the observed associations. Further details about the MMPI-2 are given in the Supplementary Text.

The primary measure of entrepreneurial behavior (Figure 1) was self-reported history of business venture creation, categorized as ‘never’, ‘once’, and ‘more than once’. In some analyses we used a dichotomized measure of entrepreneurial history (yes/no), for statistical tractability; results using the 2-level or 3-level measure of entrepreneurial history were consistent, as detailed throughout. Other aspects of entrepreneurship were assessed in the three studies using a number of well-validated instruments. In Study 1, we assessed the Business Risk Taking (BRT) scale (60), a revised version of the Entrepreneurial Intentions (EI-r) scale (61), and the Nascent Entrepreneur measure (63, 75). Details of these measures, including characteristics of the EI-r, are given in Supplementary Text. In Study 2 we used the Entrepreneurial Outcomes Inventory (76) (EO, (69)) to assess different types of entrepreneurship; details of these scales are given in Supplementary Text.

### Analysis

All data were analyzed using IBM SPSS Statistics 24.0 and R 3.5.1. De-identified data and R scripts are available at https://github.com/helenpush/HPM.git.

The main dependent variables of interest were three measures of hypomanic tendencies: HPS-s, Ma, and HPS (dependent variables, DV); these three measures correlate with one another (See Figure S1 and Supplementary Text). The independent variable for the primary analysis was history of business creation (three levels: “none created,” “one created,” “multiple created”). All measures of HPM were nearly normally distributed in all three samples (skewness was between −0.5 and 0.5); see Tables S8, S12, and S16. All three DV measures significantly correlated with age (HPS-s: r = −0.18, p < 0.001; Ma: r = −0.27, p < 0.001; HPS: r = −0.22, p< 0.001), and one differed across gender (Ma: f = 16.0, m = 18.8, F(1,421) = 7.36 p = 0.002). Age was broadly distributed in our samples (from late teens to seventies), and both age and gender have been previously shown to correlate with business creation activities (77, 78). Therefore, to examine the relation of interest while avoiding potential biases, we used age- and gender- corrected DV measures in all analyses. We ran linear regression analyses with dependent variables measures of hypomanic tendencies (HPS-s, Ma, and HPS), and independent variables age and gender. 7 respondents from Study 2 did not report their age or gender, and 7 respondents from Study 3 did not report their age or gender. Standardized residuals from these analyses were used as main dependent variables of interest for the primary analyses; residuals in all three samples were normally, or near normally (skewness was between −0.5 and 0.5) distributed. In all three samples, distributions of participants across the three groups of history of business creation were highly unbalanced (see sample characteristics in Supplementary Text), rendering the data ill-suited to analysis by ANOVA (79). Therefore, to analyze the relationship between age- and gender-corrected residuals of measures of HPM (DV) and history of business creation (IV), we used independent samples Kruskal-Wallis tests and a series of linear regressions with a heteroskedasticity-consistent variance covariance matrix (robust linear regressions (80)).

Further details and secondary analyses are described in Supplementary Text.

## Supplementary Text

### Study 1 – Design, instruments, and sample

#### Data collection and sample characteristics

A survey was sent during 2013-2014 to 12,000 household addresses in the state of Kentucky, a largely rural US state. Addresses were obtained from USADATA, covering 79 of Kentucky’s 120 counties—56 rural farming counties, 12 mining counties, and 11 urban counties, classified according to the US Department of Agriculture’s Rural-Urban Continuum Codes. For each occupation category of interest (self-employed, farming, neither self-employed nor farming) a random address list for the sample area was generated to be representative of the distribution of occupation by county. Completed surveys were returned by 1481 residents, a response rate of 12.3%; 1387 were usable for the analysis. Respondents’ ages ranged from 14 to 94 (mean = 55 ± 0.4); 60% of them were females; 52% lived in urban counties; 55.2% had no business experience, 16.9% had started a business in the past, 23% were running an established business now, and 3.5% were in the process of starting a new business. Income ranged from less than $20,000 to more than $160,000; education ranged from less than high school to a graduate degree. In addition to the HPS-s scale, the survey included a series of questions about history of business creation, current state of business activities, business risk taking, entrepreneurial intentions, and socio-demographic characteristics. This allowed us to examine the relationship between hypomanic tendencies and various measures of entrepreneurial behavior, and test whether these relationships differed across gender and rural/urban contexts. Demographic characteristics of this sample are given in Table S1. Note that nascent entrepreneurs include people with diverse business experience (see Table S2).

#### Development of the shortened hypomanic personality scale (HPS-s)

Hypomania is often measured using the hypomanic personality scale (HPS) of Eckblad and Chapman (46). The full HPS was used in Study 3, and further details are presented below. However, this scale is lengthy, and some of its items have an explicitly clinical connotation (e.g. “I am frequently so ‘hyper’ that my friends ask me what drug I’ve been taking” or “There are often times when I am so restless that it’s impossible for me to sit still”). When designing the large-scale survey instrument for Study 1, we conducted a focus group with 8 members of a Kentucky statewide entrepreneurship program to assess whether the format and framing of the HPS was appropriate (81). These experts strongly suggested that we exclude “clinical-sounding items” and shorten the scale, because our target participants would not respond to the survey otherwise. Accordingly, we modified HPS, following their suggestions. The resulting scale, the HPS-s, consists of 13 items (see Table S3). To validate the HPS-s, we collected data from 49 individuals (15 males, 39 white, ages 20-77, mean = 43±2) using the eLab service of the Yale School of Management (Yale SOM; https://elab.som.yale.edu/). The survey included both the full HPS and the HPS-s items, several demographic questions, and questions about self-employment experience. Characteristics of this sample are presented in Table S4. HPS-s strongly correlated with HPS (Spearman’s ρ = 0.72 p < 0.001, large effect size; Figure S1), as well as with each of the HPS subscales (all p < 0.01, large or medium-to-large effect sizes; see Table S4). Internal consistency of the scale in Study 1 sample was acceptable (Chronbach’s α = 0.75).

#### Business Risk Taking (BRT) Scale (60)

The BRT is an 8-item questionnaire used to access the likelihood that individuals may engage in risky business activities, such as “Borrowing 20% of annual income to invest in own business” or “Starting a new business with a family member” (see Table S5). Each is scored using a Likert scale from 1= “very unlikely” to 5 = “very likely”. Internal consistency of the scale in Study 1 sample was good (Chronbach’s α = 0.83).

#### Nascent Entrepreneur (NE) measure (63, 75)

The NE measure is a binary variable (“yes”/”no”). Individuals are identified as nascent entrepreneurs if they were in the process of starting a new business or nonprofit, and have completed at least 3 steps entailed in this process (see Table S6).

#### Entrepreneurial Intentions (EI) Scale, Revised

The individual entrepreneurial intentions scale (61) was modified based on the feedback from a focus group with 8 members of a Kentucky statewide entrepreneurship program (three questions were combined into one as repetitive and 1 question was added). The resulting scale contained 8 questions: 4 content questions and 4 distractors (Table S7), each scored using a Likert scale from 1= “very untrue” to 6 = “very true”. Internal consistency of the scale in Study 1 sample was acceptable (Chronbach’s α = 0.76).

Tables S1 and S8 report summary statistics for measures of entrepreneurship used in Study 1.

### Study 1 - Analysis

Distribution of individuals across groups with different levels of business experience was highly uneven in the Study 1 sample: 765 respondents had never started a business, 434 had started one business, and 181 had started more than one business. Variances were not homogeneous across these three groups (Levene statistics (2,1224) = 6.29 p = 0.002); studentized Breusch-Pagan test (82) for heteroskedasticity was BP(2) = 5.42 with p = 0.07, indicating a trend toward heteroskedasticity. Thus, we employed an independent samples Kruskal-Wallis test and a robust linear regression (80) to analyze the relationships between HPS-s (DV) and a history of business creation (IV; BUS_history, three levels: “none created,” “created one,” “created multiple”); see Figure 1A.

When we measured business creation at two levels rather than 3, individuals with (N = 765) and without (N = 619) business experience were approximately equally represented in the sample from Study 1. However, variances were not homogeneous across these two groups (Levene statistics (1,1382) = 5.91 p = 0.02); the studentized Breusch-Pagan test (82) for heteroskedasticity was BP(1)=3.74 with p = 0.05 could not reject the null hypothesis of homoskedasticity. Thus, we again employed an independent samples Kruskal-Wallis test and a robust linear regression (80) to analyze the relationships between HPS-s (DV) and a dichotomous measure of business entrepreneurship (IV; BUS, two levels: “never created a business,” “created a business”); see Figure S2.

Distribution of individuals across groups of different current state of business activities was highly uneven in the Study 1 sample: 765 respondents had never started a business, 229 had started a business in the past that was no longer operational, 322 respondents had started a business that continued to be operational, and 49 were currently in the process of starting a new business. Variances were not homogeneous across these four groups (Levene statistics (3,1223) = 4.20 p = 0.006); the studentized Breusch-Pagan test (82) for heteroskedasticity was BP(3) = 7.68 with p = 0.05 could not reject the null hypothesis of homoskedasticity. Therefore, we employed an independent samples Kruskal-Wallis test and a robust linear regression (80) to analyze the relationships between HPS-s (DV) and the current state of business activities (IV; BUS_current, four levels: “none created,” “created in the past, not operational,” “created in the past, operational,” “currently in the process”); see Figure 2A.

The distribution of entrepreneurial intentions (EI) in the sample was moderately positively skewed (mean = 10.2 ± 0.1, median = 9, skewness = 0.85 ± 0.07, kurtosis = 0.40 ± 0.14); with only ~8% of respondents with scores in the top 25th percentile of the scale. Therefore, to examine the relationship between HPS-s and EI, we employed a quantile regression analysis, which is robust to the influence of outliers (‘quantreg’ package in R (67)). Control variables included age and gender (since they have been shown to influence entrepreneurial intentions (77)) and business risk taking (BRT); both HPS-s and BRT were standardized; see Table S9.

### Study 2 - Design, instruments, and sample

Subjects from the general population were recruited to complete a computer-based survey through Behavioral Lab at Yale School of Management during 2016; 550 respondents participated in the study.

The survey included:

- The first 370 questions of the MMPI-2 (49); these questions include all items from the main 9 personality scales;
- A series of questions about history of business creation, current state of business activities, and the Entrepreneurial Outcomes Inventory; and
- A series of questions about socio-demographic characteristics.

MMPI-2 includes several indices designed to evaluate validity of the collected data:

- Number of missing responses;
- F-Infrequency - validity scale designed to detect over endorsement of symptoms;
- VRIN/TRIN – validity scale designed to measure random and fixed responding, respectively;
- Fp - validity scale designed to detect over endorsement of symptoms when the differential diagnosis involves genuine psychopathology.

Data validity was evaluated using these MMPI-2 indices as depicted by a Figure S3. After applying these criteria, a total of 127 respondents were excluded; in addition, 6 participants did not respond to questions about their business history, leaving 417 subjects for the planned analyses. Subject characteristics are given in Table S10.

#### The Minnesota Multiphasic Personality Inventory, MMPI-2 (49)

The MMPI-2 is used to screen for personality and psychosocial disorders in adults and adolescents (over age 14). It is also frequently administered as part of a neuropsychological test battery to evaluate cognitive and personality functioning. The nine scales of the MMPI-2, including the hypomania scale (Ma), have been designed to detect and evaluate dimensions of personality variation that are associated with psychopathology. The main criterion for acceptance or rejection of test items during scale development was their ability to differentiate between clinical (e.g. individuals with hypomania or mild mania) and normal groups (68). A subscale that evaluates individual hypomanic tendencies is detailed below. Table S11 provides a short description of the remaining MMPI-2 subscales that were used in this study.

#### Hypomania scale of the MMPI-2 (Ma)

The Ma scale is calculated from 46 items and measures clinically relevant variations in hypomanic tendencies (74). Even though the Ma scale was developed for use in clinical research and practice, it has also been used to study occupational differences (83-85), differences between prisoners and the general population (86, 87), relationships between personality types, individual traits, and individual life experiences in the general population (88-93), and age and gender related differences in individual characteristics (94). We used the Ma scale, along with other scales of MMPI-2, in Study 2. Internal consistency of the scale in Study 2 sample was acceptable (Chronbach’s α = 0.71).

85 participants (34 females) from Study 3 completed both HPS and Ma scales, allowing us to test their convergent validity. These two measures of hypomanic tendencies significantly correlated (pooled: r = 0.49 p < 0.001; females: r = 0.55 p = 0.01; males: r = 0.50 p < 0.001, large effect sizes).

#### Entrepreneurial outcomes inventory (EO, (69))

The EO is a 31-item questionnaire designed to access diversity of entrepreneurial activities and outcomes, including business entrepreneurship (“started own business”), student entrepreneurship (e.g. “initiated/created student organization”), corporate entrepreneurship (e.g. “solved a longstanding organizational problem”), and invention entrepreneurship (e.g. “published or patented an invention or original piece of work”).

Tables S13 reports summary statistics of these measures of entrepreneurship in Study 2.

### Study 2 - Analysis

Scores on two MMPI scales were moderately positively skewed (Hs and Sc), as displayed in Table S12. Scores on the remaining six MMPI-2 scales were normally or near normally distributed in our sample (skewness was between −0.5 and 0.5; D, Hy, Pd, Pa, Pt, Si). Scores on five of these scales significantly negatively correlated with age and on four of these scales were significantly different across genders; see Table S12. We ran a series of linear regression analyses with scores on these MMPI-2 scales as dependent variables measures, and age and gender as independent variables. Standardized residuals from these analyses were used as age- and gender-corrected measures of individual differences. Two of these residuals were moderately positively skewed (Hs: skewness = 0.97; Sc: skewness = 0.53); the remaining measures were near normally distributed (skewness between −0.5 and 0.5). To analyze the relationships between age and gender corrected MMPI-2 measures of personality characteristics (DVs) and business entrepreneurship, we employed Mann-Whitney U test (IV: BUS) and a series of robust linear regressions (IV: BUS_history) (79, 95). Since 9 identical tests were conducted (on 9 MMPI-2 subscales), we corrected the significance level for multiple comparisons (p < (0.05/9) = 0.006).

Since measures of student, corporate, and invention entrepreneurship were positively skewed (Table S13), we employed Spearman’s correlation analyses to examine the relationship between these measures of entrepreneurial outcomes and MMPI-2 measures of personality. Since 27 identical tests were conducted (on 9 MMPI-2 subscales vs. 3 EO measures), we corrected the significance level for multiple comparisons (p < (0.05/27) = 0.002).

There were no significant associations between MMPI dimensions other than Ma and entrepreneurship, either when measured at three levels as in Figure 1 (shown in Figure S14, below) or when dichotomized (shown in the main text, Table 1).

### Study 3 - Design, instruments, and sample

216 current or past students of MBA programs were recruited through professors at several U.S. and European universities during 2016. Respondents completed a Qualtrics-based online survey, which included the full HPS scale and a series of questions about history of business creation, current state of business activities, current work status, the Entrepreneurial Outcomes Inventory, and questions about personality and socio-demographic characteristics. An additional 219 current or past students of MBA programs were recruited through the Max Planck Institute for Innovation and Competition during 2017; these respondents completed a shorter version of the survey to further increase the sample. Both surveys were in English. Data from both waves were pooled; 389 responses were usable to analyze relationships between hypomanic tendencies and entrepreneurial behavior; 108 responses were usable to analyze relationships between hypomanic tendencies and entrepreneurial success.

Respondents’ ages ranged from 19 to 77 (median = 36); 28% of them were females; 47.5% had no business experience, 32.8% have started one business, 19.7% started multiple businesses. Income ranged from less than $5,000 to more than $300,000 (median = $85,000). 45% of respondents were from France, 30% were from Germany, 10% were from the U.S., the remaining participants were from other mostly European countries. Of those, who started their own business, 12% had their business in operation less than 1 year, 29% - 1-3 years, 22% - 3-7 years, and 37% for more than 7 years.

#### Hypomanic personality scale (HPS)

In this study we used the full HPS, rather than the shortened version used in Study 1. The HPS is a 48-items questionnaire that was originally designed to evaluate the risk for development of bipolar disorder in individuals from the general population. It has been suggested to “identify persons with hypomanic personality, an overactive, gregarious style in which episodes of hypomanic euphoria are likely to occur” (Eckblad and Chapman, 1986, p 214). Sample items include the following: “Sometimes ideas and insights come to me so fast that I cannot express them all” (keyed true), and “I would rather be an ordinary success in life than a spectacular failure” (keyed false). The scale has good internal and test-retest reliability (46). More recent research argued that HPS “is unlikely to simply be a measure of personality style and appears strongly confounded by hypomanic/manic mood symptoms” ((66) p. 654). Another line of recent research examined multi-dimensionality of HPS (96-98). A three-factor solution suggested social vitality (SV), mood volatility (MV), and excitement (Ex) subscales (70).

HPS has been shown to be predictive of a risk for developing bipolar disorder and several other forms of psychopathology (99). Hypomanic tendencies (measured by HPS) have been shown to be heritable and related to a number of polymorphisms in genes associated with dopamine neurotransmission and metabolism (72). The aggregate cross-national lifetime prevalence of BP-I disorder is 0.6% (1.0% for the U.S.), BP-II is 0.4% (1.1% for the U.S.), subthreshold BP is 1.4% (2.4% for the U.S.), and Bipolar Spectrum (BPS) is 2.4% (4.4% for the U.S.). In contrast, according to several short-term U.S. longitudinal studies, approximately 25%-29% of individuals who score in the upper quantile of HPS (> 30) develop bipolar disorder within 3 years (100, 101); up to 58% of these individuals develop a bipolar spectrum disorder within 3 years (100). It is noteworthy that approximately half of these high-scoring individuals do not develop psychopathology, at least within a few years (100). Despite this, high HPS is often considered a risk factor for the development of clinical hypomania or mania (29, 102, 103). Some research suggests a more nuanced relationship. For instance, a longitudinal 13-year follow-up study found that only individuals with high scores on both the HPS and the Impulsive-Nonconformity Scale (104) experienced greater rates of bipolar mood disorders, borderline personality symptoms, poorer overall adjustment, and higher rates of arrest than the remaining HYP or control participants (105).

The HPS has recently been applied to the study of entrepreneurship, with somewhat mixed results. For instance, high HPS scores were noted more frequently among business founders (61%) than among the employed executives (43%) or other professionals (20%) (32). Many personality traits tied to high HPS scores appear to be related to entrepreneurial intent and entry (10). On the other hand, in another study, HPS did not predict entrepreneurial intentions among college students and was not tied to selected indices of entrepreneurial success among entrepreneurs (33).

We use the full HPS to quantify hypomanic tendencies in Study 3. Internal consistency of the scale and each of the subscales were acceptable/good in Study 3 sample (HPS: Chronbach’s α = 0.87, SV: Chronbach’s α = 0.77, MV: Chronbach’s α = 0.80; Ex: Chronbach’s α = 0.71).

#### Entrepreneurial Success

Respondents who had started their own business were asked to report annual revenue of their most successful business at its peak, in U.S. dollars.

Table S16 reports summary statistics for measures of hypomanic tendencies and entrepreneurship, Study 3.

### Study 3 - Analysis

Two subscales of HPS, SV and MV, were slightly skewed (skewness was between 0 and 0.65); EX was somewhat more skewed (but still less than 1); see Table S16. Scores on these subscales significantly correlated (Spearman’s ρ > 0.48; p<0.001, large effect size), reflecting that these distinct traits interrelate to constitute hypomanic tendencies. We ran linear regression analyses with scores on the HPS subscales as dependent variables measures and age and gender as independent variables. Standardized residuals from these analyses were used as age and gender-corrected measures of individual differences. We employed Kruskal-Wallis tests and a series of robust linear regressions (79, 95) to examine how scores on these subscales (DVs) varies among individuals with different histories of business creation (IV; see main text, Table 2).

A subset of Study 3 participants (N = 214) responded to the Entrepreneurial Outcomes questionnaire (EO). As in Study 2, measures of student, corporate, and invention entrepreneurship were positively skewed; see Table S16. The relationship of both the full HPS and its subscales with different types of entrepreneurship, as defined by the EO, was less strong than with history of business entrepreneurship in this sample, consistent with our findings in an independent sample in Study 2 (see Table S17; compare to main text, Table 1).

A subset of Study 3 participants reported annual revenue of their most successful business (N = 109). This measure was not normally distributed in our sample (Best Annual Revenue: min = $0, max = $1B, mean = $12,809,079 ± $9,262,190, median = $150,000, see Table S16), with only 4% of individuals reporting revenue higher than the sample average. To analyze the relationship between the best annual revenue and HPS scale and subscales (age and gender corrected), we employed quantile regressions, which are robust to effects of outliers (‘quantreg’ package in R (67)). The extreme quantiles (<0.20 and > 0.80) had unreliable 95% confidence intervals for our sample sizes (106), and are not reported. Total HPS scale was not associated with entrepreneurial success in any of the quantiles. For association of HPS subscales with entrepreneurial success see Figure 3 in the main text and Table S18.

**Figure S1.**
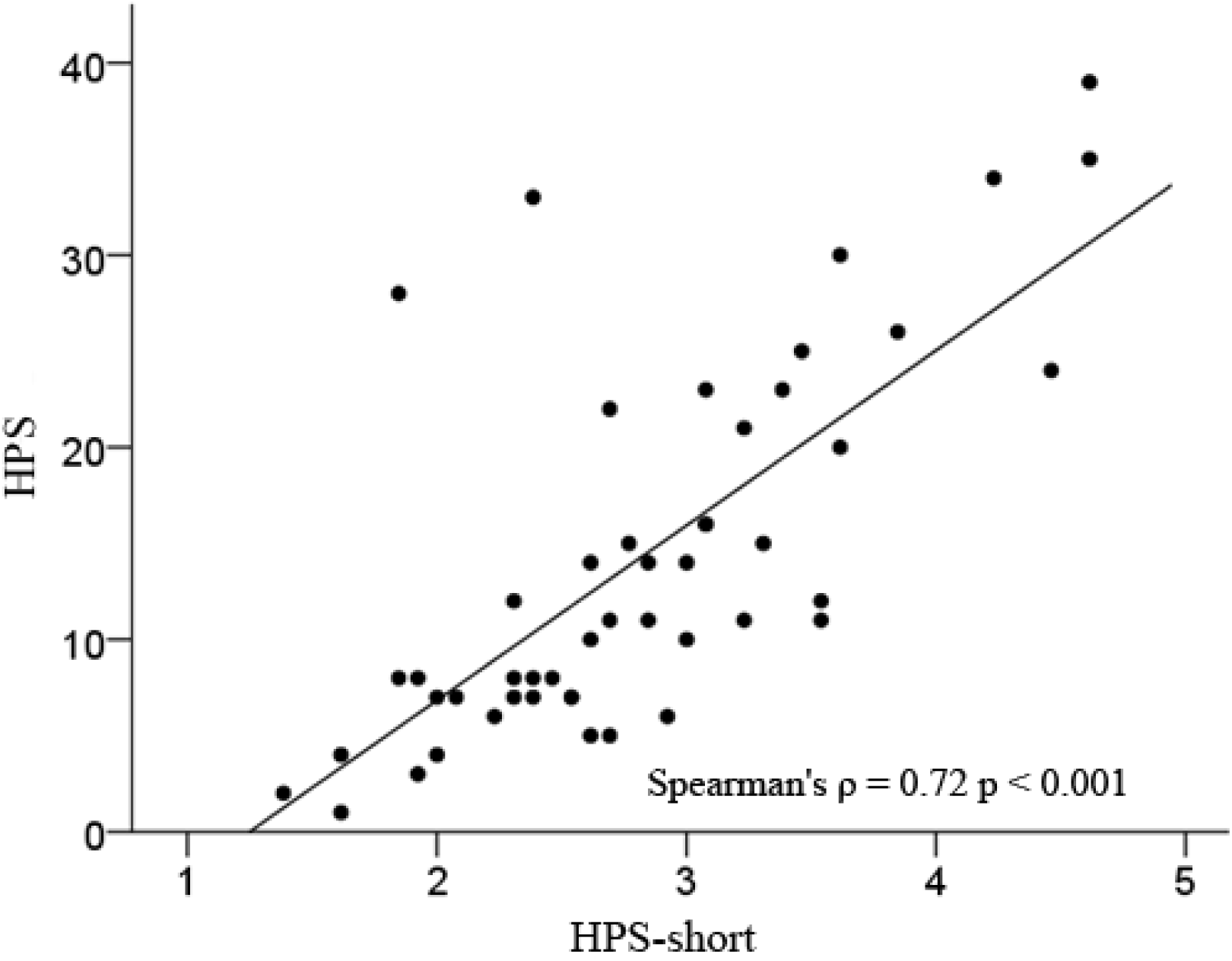
HPS and HPS-s tightly correlated in a convenience sample of 49 individuals.

**Figure S2.**
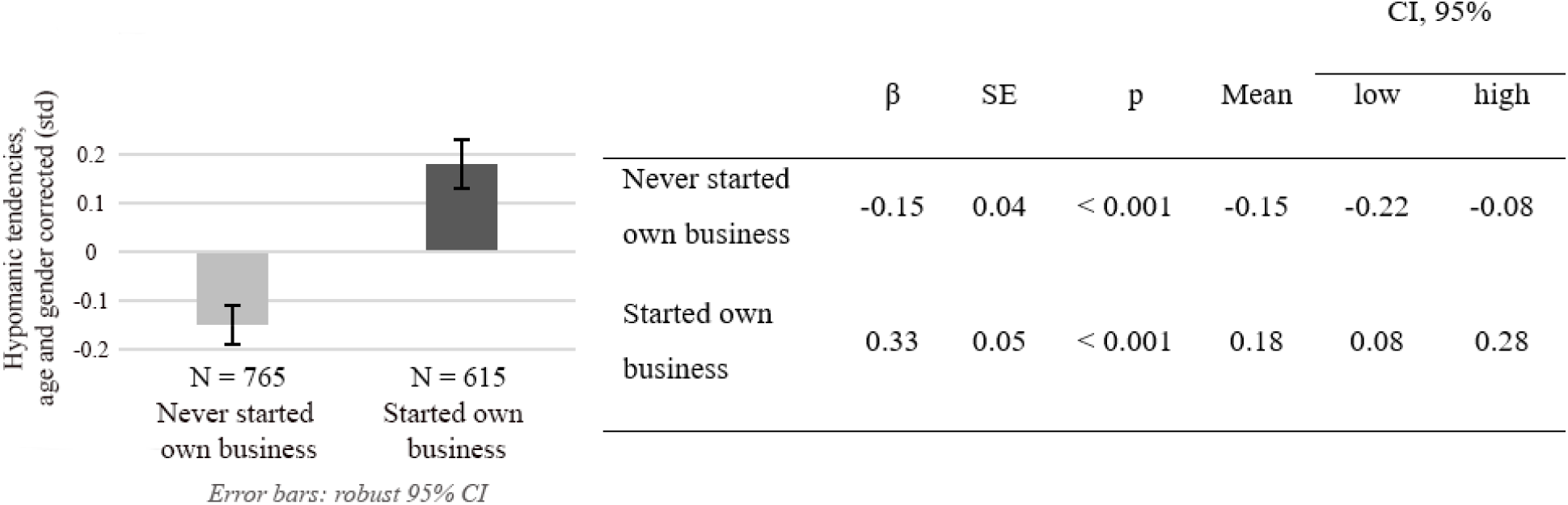
Relationship between dichotomized history of entrepreneurship and hypomania (HPS-s) in Study 1.

**Figure S3.**
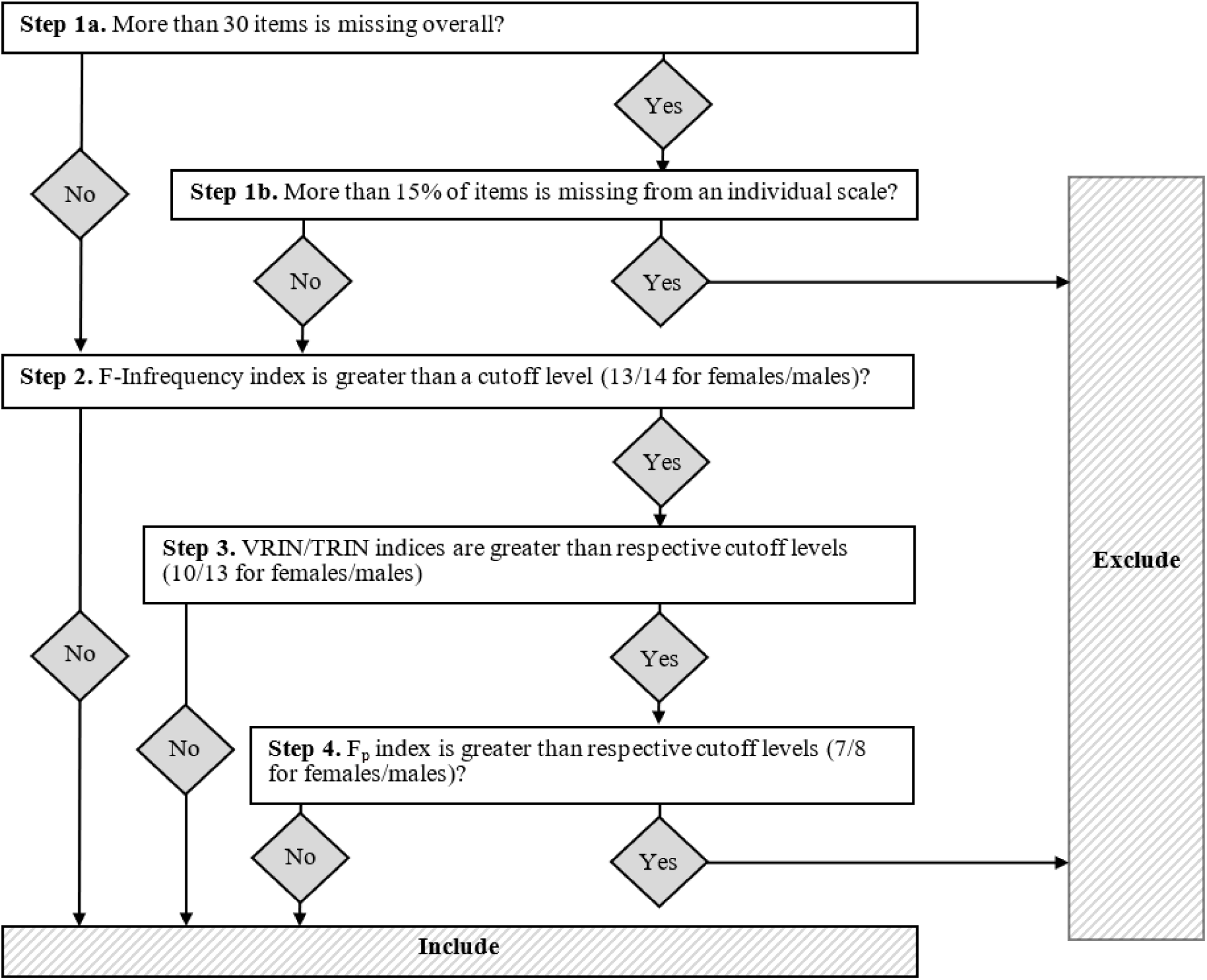
Data validation steps for Study 2.

**Table S1.**
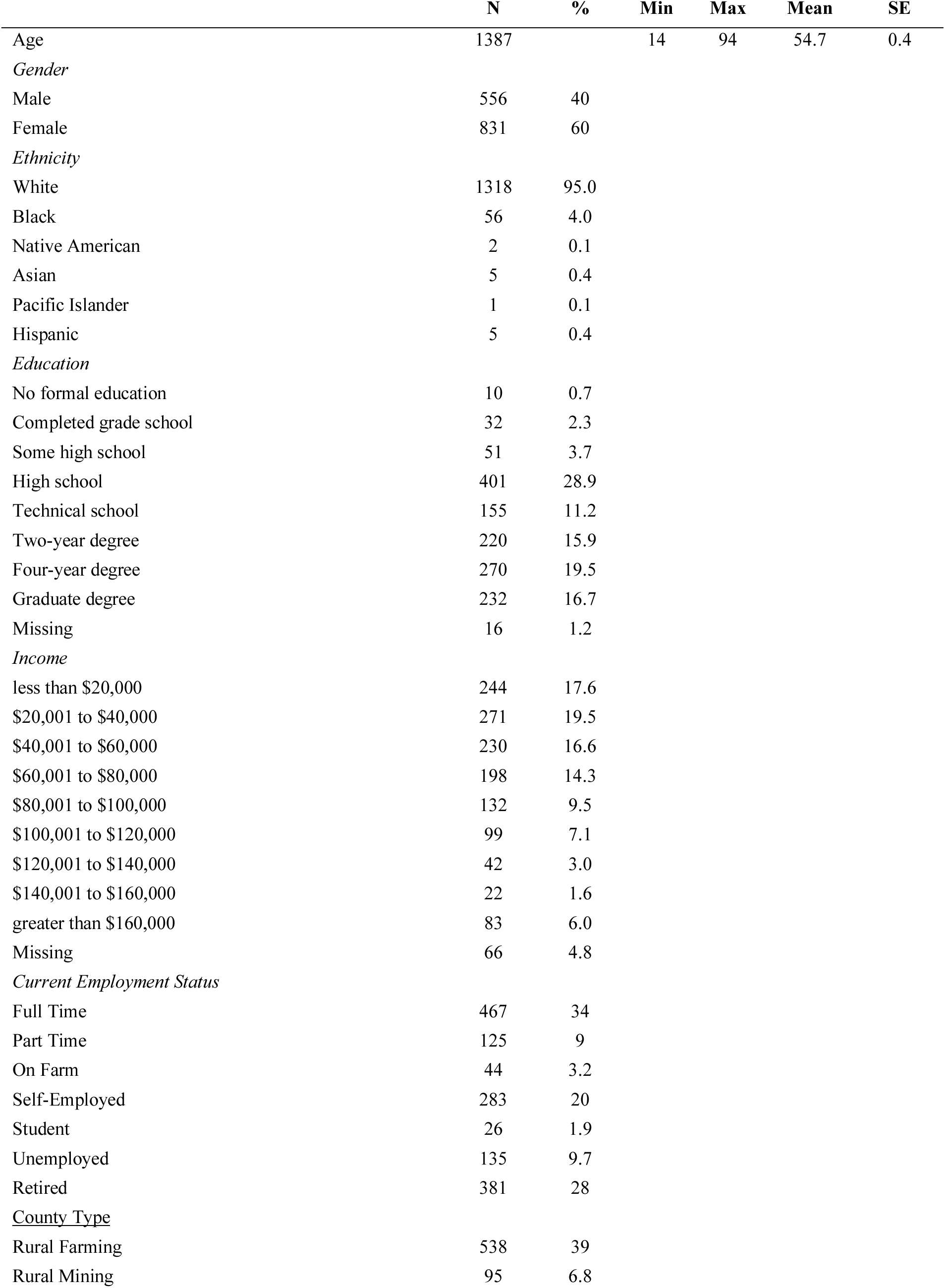
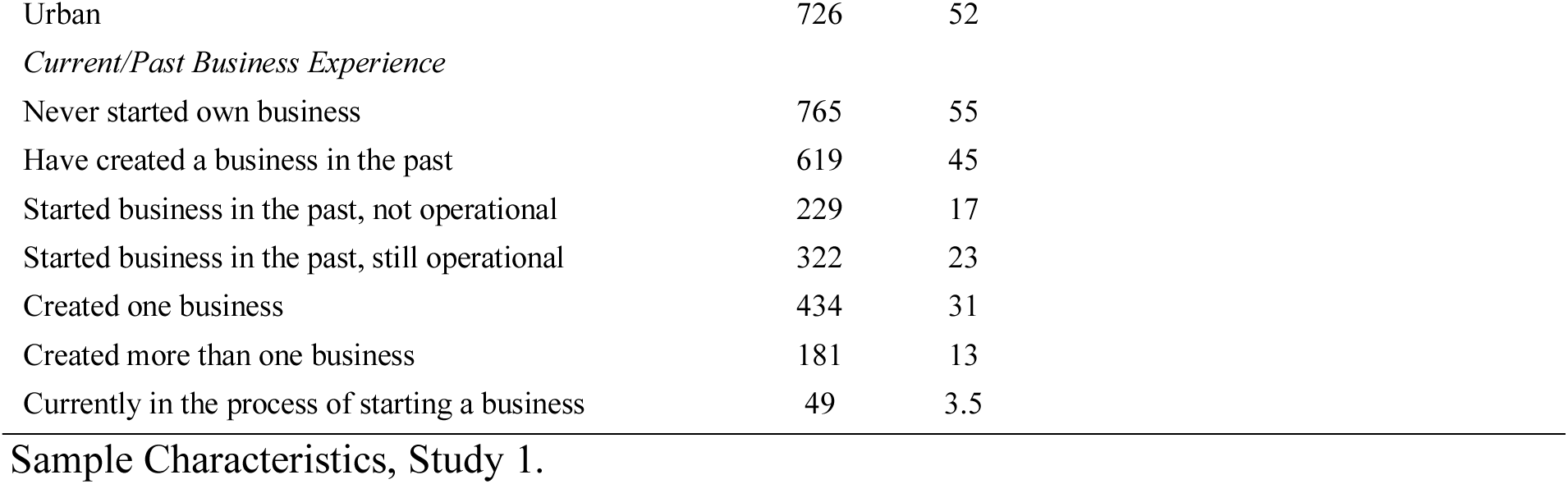

|  | **N** | **%** | **Min** | **Max** | **Mean** | **SE** |
| --- | --- | --- | --- | --- | --- | --- |
| Age | 1387 |  | 14 | 94 | 54.7 | 0.4 |
| *Gender* |  |  |  |  |  |  |
| Male | 556 | 40 |  |  |  |  |
| Female | 831 | 60 |  |  |  |  |
| *Ethnicity* |  |  |  |  |  |  |
| White | 1318 | 95.0 |  |  |  |  |
| Black | 56 | 4.0 |  |  |  |  |
| Native American | 2 | 0.1 |  |  |  |  |
| Asian | 5 | 0.4 |  |  |  |  |
| Pacific Islander | 1 | 0.1 |  |  |  |  |
| Hispanic | 5 | 0.4 |  |  |  |  |
| *Education* |  |  |  |  |  |  |
| No formal education | 10 | 0.7 |  |  |  |  |
| Completed grade school | 32 | 2.3 |  |  |  |  |
| Some high school | 51 | 3.7 |  |  |  |  |
| High school | 401 | 28.9 |  |  |  |  |
| Technical school | 155 | 11.2 |  |  |  |  |
| Two-year degree | 220 | 15.9 |  |  |  |  |
| Four-year degree | 270 | 19.5 |  |  |  |  |
| Graduate degree | 232 | 16.7 |  |  |  |  |
| Missing | 16 | 1.2 |  |  |  |  |
| *Income* |  |  |  |  |  |  |
| less than \$20,000 | 244 | 17.6 | | | | |
| \$20,001 to \$40,000 | 271 | 19.5 | | | | |
| \$40,001 to \$60,000 | 230 | 16.6 | | | | |
| \$60,001 to \$80,000 | 198 | 14.3 | | | | |
| \$80,001 to \$100,000 | 132 | 9.5 | | | | |
| \$100,001 to \$120,000 | 99 | 7.1 | | | | |
| \$120,001 to \$140,000 | 42 | 3.0 | | | | |
| \$140,001 to \$160,000 | 22 | 1.6 | | | | |
| greater than \$160,000 | 83 | 6.0 | | | | |
| Missing | 66 | 4.8 |  |  |  |  |
| *Current Employment Status* |  |  |  |  |  |  |
| Full Time | 467 | 34 |  |  |  |  |
| Part Time | 125 | 9 |  |  |  |  |
| On Farm | 44 | 3.2 |  |  |  |  |
| Self-Employed | 283 | 20 |  |  |  |  |
| Student | 26 | 1.9 |  |  |  |  |
| Unemployed | 135 | 9.7 |  |  |  |  |
| Retired | 381 | 28 |  |  |  |  |
| <u>County Type</u> |  |  |  |  |  |  |
| Rural Farming | 538 | 39 |  |  |  |  |
| Rural Mining | 95 | 6.8 |  |  |  |  |

|  |  |  |
| --- | --- | --- |
| Urban | 726 | 52 |
| *Current/Past Business Experience* |  |  |
| Never started own business | 765 | 55 |
| Have created a business in the past | 619 | 45 |
| Started business in the past, not operational | 229 | 17 |
| Started business in the past, still operational | 322 | 23 |
| Created one business | 434 | 31 |
| Created more than one business | 181 | 13 |
| Currently in the process of starting a business | 49 | 3.5 |

Sample Characteristics, Study 1.

**Table S2.**

|  | History of Business Creation |  |  | Total |
| --- | --- | --- | --- | --- |
|  | None created | One created | Many created - Serial entrepreneur |  |
| Currently not starting a business | 757 | 408 | 158 | 1323 |
| Currently starting a business - Nascent Entrepreneur | 8 | 26 | 23 | 57 |
| Total | 765 | 434 | 181 | 1380 |
| Missing |  |  |  | 7 |

Serial Entrepreneurs and Nascent Entrepreneurs, Study 1.

**Table S3.**

| # | item |
| --- | --- |
| 1. | I do most of my best work during brief periods of intense inspiration |
| 2. | I tend to get very passionate about things I care about |
| 3. | I am frequently in such high spirits that I can't concentrate on any one thing for too long |
| 4. | I frequently find that my thoughts are racing |
| 5. | When with groups of people I usually prefer to let someone else be the center of attention (R) |
| 6. | I am a very even-tempered person (R) |
| 7. | Sometimes ideas and insights come to me so fast that cannot express them all |
| 8. | I often get contagiously happy and energetic |
| 9. | When I work on an important project, I am able to work for a long time non-stop, without much sleep |
| 10. | People often come to me when they need a clever idea |
| 11. | I frequently write down the thoughts and insights that come to me when I am thinking especially creatively |
| 12. | I have an uncommon ability to persuade and inspire others |
| 13. | I am very good in a "crisis mode," but I prefer to delegate routine tasks to somebody else |

Short version of a hypomanic personality scale. The scale score is an average of scores across all items; R – this item is reversed scored.

**Table S4.**
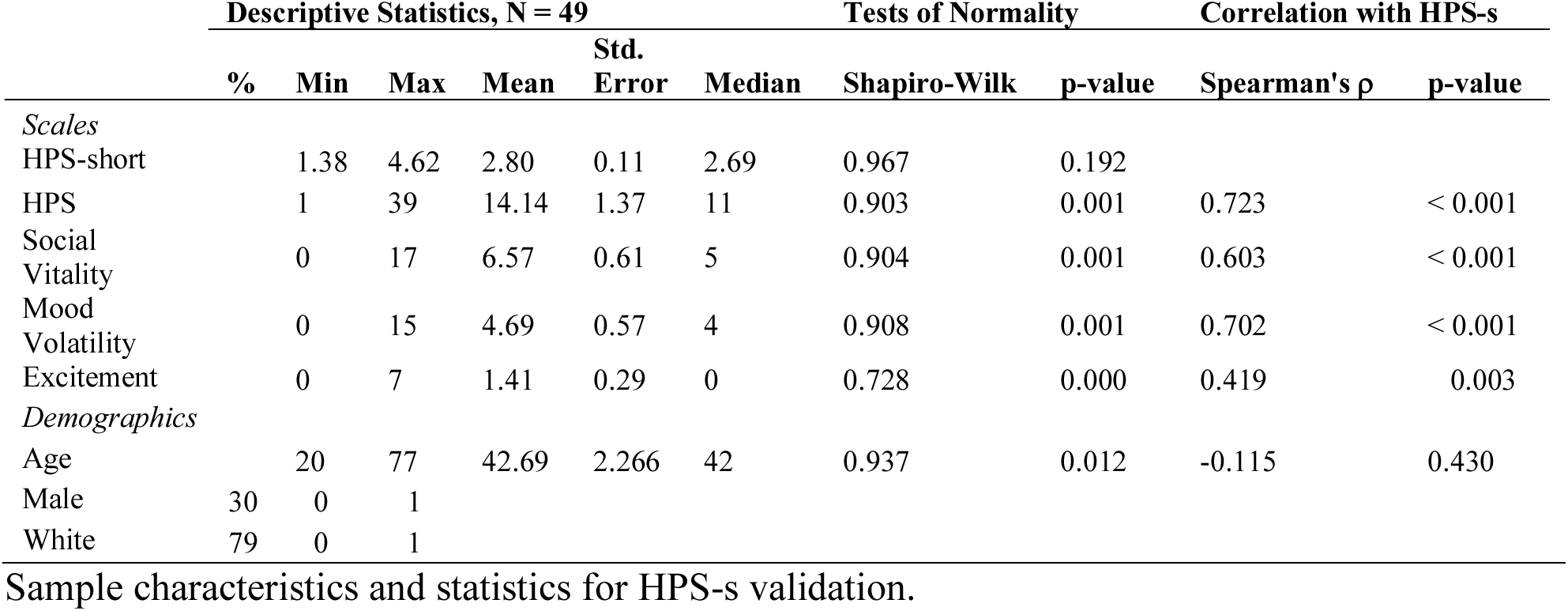

**Table S5.**

|  |  |
| --- | --- |
| BRT.1 | Borrowing 20% of your annual income from a bank to invest in your own business |
| BRT.2 | Quitting a job that pays you \$60,000 a year to start your own business |
| BRT.3 | Taking an equity loan on your main residence to invest money in your own business |
| BRT.4 | Starting a new business alone |
| BRT.5 | Starting a new business with a family member |
| BRT.6 | Starting a new business with a partner(s) other than family members |
| BRT.7 | Starting a new business when other employment opportunities are not available |
| BRT.8 | Starting a new business while maintaining other employment |

**Table S6.**

|  |  |
| --- | --- |
| NE.2.1 | Researched the business idea |
| NE.2.2 | Started networking |
| NE.2.3 | Wrote a business plan |
| NE.2.4 | Sought out mentoring/assistance |
| NE.2.5 | Pursued financing |
| NE.2.6 | Started work on production |
| NE.2.7 | Did a trial run |
| NE.2.8 | Worked on marketing |

Nascent Entrepreneur questionnaire.

**Table S7.**

| Please indicate how untrue or true each of the following statements are as they pertain to you, using the following scale: 1 = Very Untrue, 2 = Untrue, 3 = Slightly Untrue, 4 = Slightly True, 5 = True, 6 = Very True |  |
| --- | --- |
| # | item |
| 1. | I intend to start a new business sometime in the future |
| 2. | I plan my future carefully (D) |
| 3. | I read business-related newspapers and books (D) |
| 4. | I never look for business start-up opportunities (R) |
| 5. | I am saving money to start a business |
| 6. | I plan my finances carefully (D) |
| 7. | I spend time learning about how to start a business |
| 8. | I often identify business opportunities for my acquaintances (D) |

Entrepreneurial Intentions scale, revised. The scale score is a sum of scores across 4 items; R – this item is reversed scored, D – this is a distraction item.

**Table S8.**
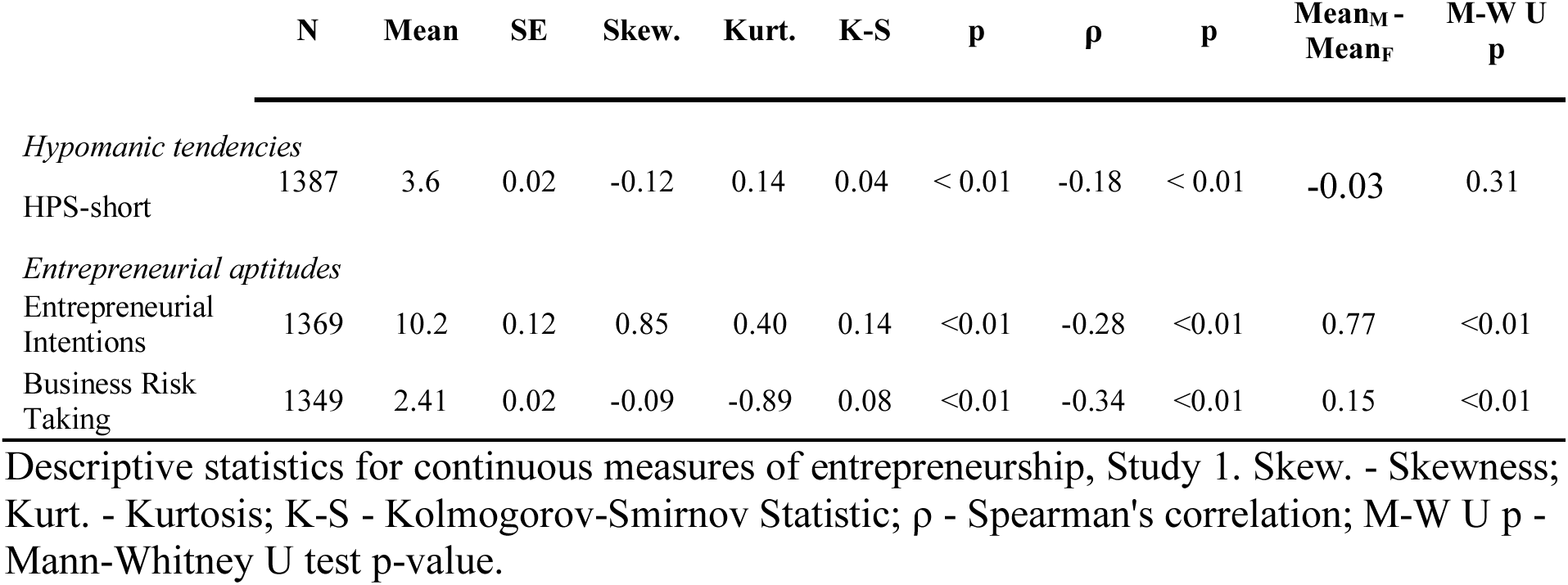

**Table S9.**

|  |  | **Intercept** | **Age** | **Male** | **HPS-s, std.** | **BRT, std.** |
| --- | --- | --- | --- | --- | --- | --- |
| 10% | $\beta$ (SE) | 5.81 (0.28) | -0.01 (0.004) | 0.31 (0.14) | 0.50 (0.07) | 0.61 (0.11) |
|  | p-value | <0.001 | 0.013 | 0.025 | <0.001 | <0.001 |
| 20% | $\beta$ (SE) | 7.92 (0.57) | -0.02 (0.01) | 0.29 (0.26) | 0.55 (0.12) | 1.10 (0.15) |
|  | p-value | <0.001 | 0.038 | 0.26 | <0.001 | <0.001 |
| 30% | $\beta$ (SE) | 9.86 (0.55) | -0.04 (0.01) | 0.38 (0.24) | 0.79 (0.11) | 1.12 (0.12) |
|  | p-value | <0.001 | <0.001 | 0.122 | <0.001 | <0.001 |
| 40% | $\beta$ (SE) | 11.12 (0.54) | -0.04 (0.01) | 0.63 (0.27) | 0.88 (0.14) | 1.01 (0.14) |
|  | p-value | <0.001 | <0.001 | 0.022 | <0.001 | <0.001 |
| 50% | $\beta$ (SE) | 11.89 (0.48) | -0.04 (0.01) | 0.59 (0.24) | 0.93 (0.12) | 1.08 (0.13) |
|  | p-value | <0.001 | <0.001 | 0.015 | <0.001 | <0.001 |
| 60% | $\beta$ (SE) | 12.67 (0.50) | -0.04 (0.01) | 0.63 (0.25) | 0.94 (0.12) | 1.21 (0.13) |
|  | p-value | <0.001 | <0.001 | 0.013 | <0.001 | <0.001 |
| 70% | $\beta$ (SE) | 13.83 (0.65) | -0.04 (0.01) | 0.95 (0.32) | 1.06 (0.16) | 1.45 (0.17) |
|  | p-value | <0.001 | <0.001 | 0.003 | <0.001 | <0.001 |
| 80% | $\beta$ (SE) | 15.34 (0.63) | -0.04 (0.01) | 0.94 (0.31) | 1.04 (0.14) | 1.93 (0.18) |
|  | p-value | <0.001 | <0.001 | 0.002 | <0.001 | <0.001 |
| 90% | $\beta$ (SE) | 17.03 (0.81) | -0.04 (0.01) | 0.98 (0.41) | 1.27 (0.21) | 2.38 (0.21) |
|  | p-value | <0.001 | 0.005 | 0.016 | <0.001 | <0.001 |

Quantile regression of the relationship between entrepreneurial intentions (EI-r) and hypomania (HPS-s).

**Table S10.**

|  | **N** | **%** | **Min** | **Max** | **Mean** | **SE** |
| --- | --- | --- | --- | --- | --- | --- |
| Age | 417 |  | 18 | 77 | 34 | 0.7 |
| *Gender* |  |  |  |  |  |  |
| Male | 129 | 30.9 |  |  |  |  |
| Female | 288 | 69.1 |  |  |  |  |
| *Ethnicity* |  |  |  |  |  |  |
| White | 321 | 77 |  |  |  |  |
| Black | 23 | 5.5 |  |  |  |  |
| Indian American | 4 | 1 |  |  |  |  |
| Asian | 59 | 14.1 |  |  |  |  |
| Islander | 3 | 0.7 |  |  |  |  |
| Hispanic | 24 | 5.8 |  |  |  |  |
| *Income* |  |  |  |  |  |  |
| less than \$5,000 | 105 | 25.2 | | | | |
| \$5,000 - \$14,999 | 34 | 8.2 | | | | |
| \$15,000 - \$19,999 | 16 | 3.8 | | | | |
| \$20,000 - \$24,999 | 11 | 2.6 | | | | |
| \$25,000 - \$29,999 | 42 | 10.1 | | | | |
| \$30,000 - \$39,999 | 36 | 8.6 | | | | |
| \$40,000 - \$49,999 | 35 | 8.4 | | | | |
| \$50,000 - \$59,999 | 28 | 6.7 | | | | |
| \$60,000 - \$69,999 | 24 | 5.8 | | | | |
| \$70,000 - \$79,999 | 11 | 2.6 | | | | |
| \$80,000 - \$89,999 | 10 | 2.4 | | | | |
| \$90,000 - \$99,999 | 14 | 3.4 | | | | |
| \$100,000 - \$149,999 | 24 | 5.8 | | | | |
| \$150,000 - \$199,999 | 10 | 2.4 | | | | |
| \$200,000 - \$300,000 | 4 | 1 | | | | |
| over \$300,000 | 6 | 1.4 | | | | |
| Missing | 7 | 1.7 |  |  |  |  |
| *Current Employment Status* |  |  |  |  |  |  |
| Full Time | 169 | 59 |  |  |  |  |
| Part Time | 77 | 18.5 |  |  |  |  |
| Self-Employed | 38 | 9.1 |  |  |  |  |
| Student | 128 | 30.7 |  |  |  |  |
| Unemployed | 49 | 11.8 |  |  |  |  |
| Other | 25 | 6 |  |  |  |  |
| *Business Experience* |  |  |  |  |  |  |
| No business experience | 345 | 82.7 |  |  |  |  |
| Started one business | 35 | 8.4 |  |  |  |  |
| Started multiple businesses | 37 | 8.9 |  |  |  |  |

Sample Characteristics, Study 2.

**Table S11.**

| **Scale** |  | **Tendencies among high scorers** |
| --- | --- | --- |
| **Hs** | 32 items, Hathaway & McKinley (1940) | Preoccupation about one's health, a tendency to exaggerate physical symptoms, demanding, whiny, immature. |
| **D** | 57 items, Hathaway & McKinley (1942) | Depression, pessimism, guilt feelings. |
| **Hy** | 60 items, Hathaway & McKinley (1944) | Somatization, conversion reactions, denial, naiveté and manipulateness. |
| **Pd** | 50 items, Hathaway & McKinley (1940) | General social maladjustment and the absence of strongly pleasant experiences. |
| **Pa** | 40 items Hathaway & McKinley (1956) | Suspicious, hostile, overly sensitive, ideas of reference, delusions of persecution. |
| **Pt** | 48 items, Hathaway & McKinley (1942) | Obsessive-compulsive anxiety, tendency towards phobias, irrational fears, lack of self-confidence, perfectionistic and dependent. |
| **Sc** | 78 items, Hathaway & McKinley (1956). | Immature/disorganized thinking, breakdown of reality testing, schizoid trends, alienation, over-investment in fantasy, non-conforming. |
| **Si** | 69 items, Drake (1946). | Timid, shy, hard to get to know, overly-sensitive, over-controlled, submissive, conventional, cautious, rigid and moody. |

Short descriptions of the remaining MMPI-2 scales.

**Table S12.**
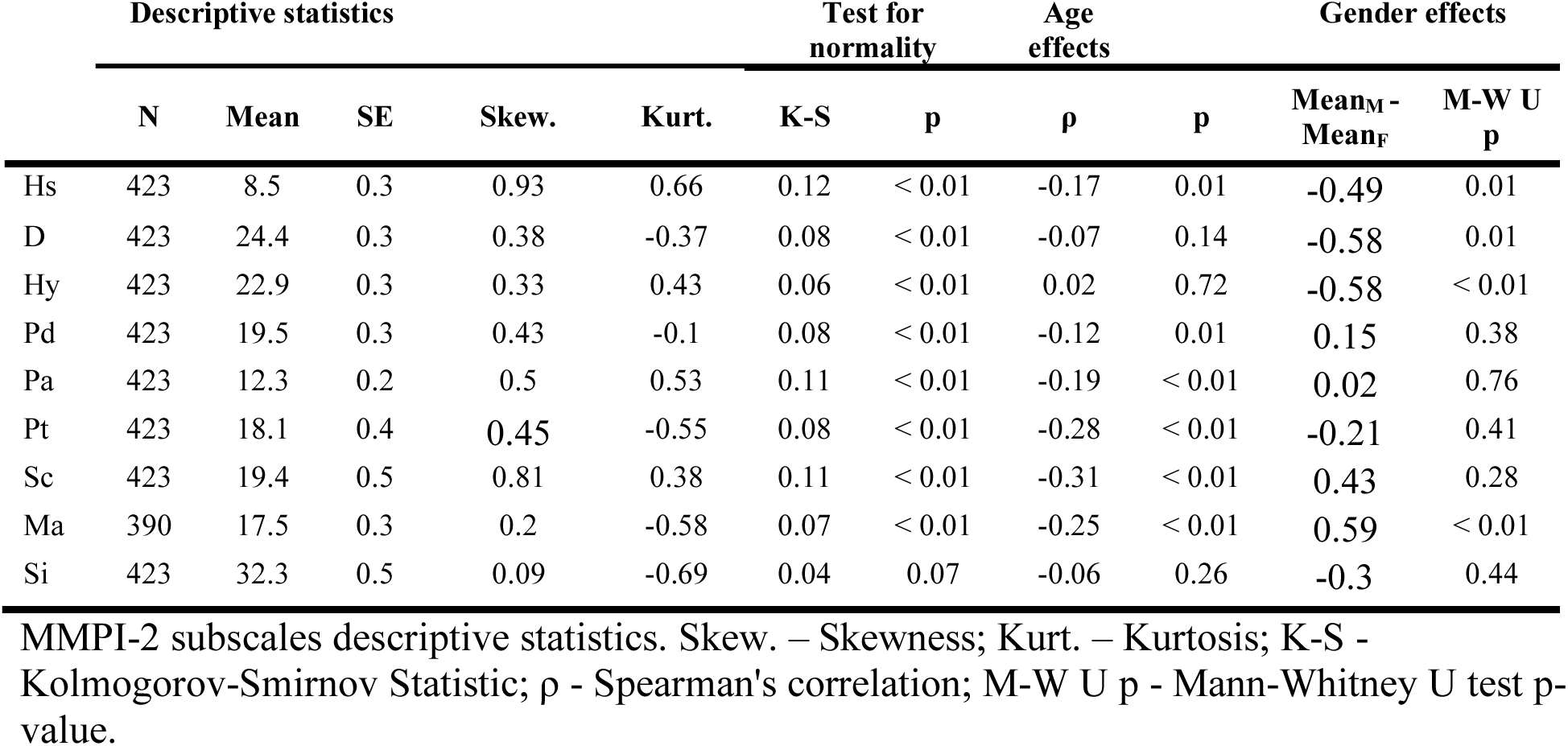

**Table S13.**

|  | Descriptive Statistics |  |  |  |  | Test for normality |  | Age effects |  | Gender effects |  |
| --- | --- | --- | --- | --- | --- | --- | --- | --- | --- | --- | --- |
| | N | Mean | SE | Skew. | Kurt. | K-S | p | $\rho$ | p | Mean<sub>M</sub> - Mean<sub>F</sub> | M-W U p |
| Student Entrepreneurship | 416 | 7.9 | 1.1 | 6.41 | 49.11 | 0.36 | < 0.01 | -0.24 | < 0.01 | -0.57 | 0.80 |
| Corporate Entrepreneurship | 416 | 25.7 | 7.0 | 14.63 | 237.76 | 0.43 | < 0.01 | 0.15 | < 0.01 | 3.69 | 0.72 |
| Invention Entrepreneurship | 416 | 18.4 | 8.2 | 11.79 | 147.50 | 0.48 | < 0.01 | <0.01 | 0.88 | -16.41 | 0.02 |

Descriptive statistics for continuous measures of entrepreneurship, Study 2. Skew. – Skewness; Kurt. – Kurtosis; K-S - Kolmogorov-Smirnov Statistic; $\rho$ - Spearman's correlation; M-W U p - Mann-Whitney U test p-value.

**Table S14.**

| | $\beta$ | SE | p | Mean | CI, 95% | |
| --- | --- | --- | --- | --- | --- | --- |
|  |  |  |  |  | low | High |
| *DV = Hs, age and gender corrected (standardized)* |  |  |  |  |  |  |
| Never started own business | -0.03 | 0.05 | 0.545 | -0.03 | -0.12 | 0.06 |
| Started one business | 0.31 | 0.21 | 0.143 | 0.28 | -0.08 | 0.63 |
| Started multiple businesses | 0.02 | 0.17 | 0.883 | -0.01 | -0.29 | 0.28 |
| *DV = D, age and gender corrected (standardized)* |  |  |  |  |  |  |
| Never started own business | 0.02 | 0.05 | 0.778 | 0.02 | -0.07 | 0.10 |
| Started one business | 0.10 | 0.16 | 0.522 | 0.12 | -0.16 | 0.40 |
| Started multiple businesses | -0.38 | 0.18 | 0.035 | -0.37 | -0.67 | -0.06 |
| *DV = Hy, age and gender corrected (standardized)* |  |  |  |  |  |  |
| Never started own business | -0.01 | 0.05 | 0.796 | -0.01 | -0.10 | 0.08 |
| Started one business | 0.19 | 0.17 | 0.255 | 0.17 | -0.11 | 0.45 |
| Started multiple businesses | -0.05 | 0.18 | 0.775 | -0.06 | -0.36 | 0.23 |
| *DV = Pd, age and gender corrected (standardized)* |  |  |  |  |  |  |
| Never started own business | 0.00 | 0.06 | 0.999 | 0.00 | -0.09 | 0.09 |
| Started one business | 0.09 | 0.18 | 0.603 | 0.09 | -0.21 | 0.39 |
| Started multiple businesses | -0.11 | 0.13 | 0.408 | -0.11 | -0.32 | 0.11 |
| *DV = Pa, age and gender corrected (standardized)* |  |  |  |  |  |  |
| Never started own business | 0.00 | 0.06 | 0.939 | 0.00 | -0.09 | 0.10 |
| Started one business | 0.12 | 0.16 | 0.473 | 0.12 | -0.15 | 0.40 |
| Started multiple businesses | -0.17 | 0.17 | 0.309 | -0.17 | -0.45 | 0.12 |
| *DV = Pt, age and gender corrected (standardized)* |  |  |  |  |  |  |
| Never started own business | -0.01 | 0.05 | 0.892 | -0.01 | -0.10 | 0.08 |
| Started one business | 0.25 | 0.20 | 0.213 | 0.24 | -0.10 | 0.57 |
| Started multiple businesses | -0.20 | 0.17 | 0.250 | -0.21 | -0.50 | 0.09 |
| *DV = Sc, age and gender corrected (standardized)* |  |  |  |  |  |  |
| Never started own business | -0.03 | 0.05 | 0.516 | -0.03 | -0.12 | 0.05 |
| Started one business | 0.31 | 0.23 | 0.176 | 0.28 | -0.11 | 0.67 |
| Started multiple businesses | 0.06 | 0.17 | 0.720 | 0.03 | -0.26 | 0.31 |
| *DV = Ma, age and gender corrected (standardized)* |  |  |  |  |  |  |
| Never started own business | -0.04 | 0.05 | 0.420 | -0.04 | -0.14 | 0.06 |
| Started one business | 0.08 | 0.17 | 0.660 | 0.04 | -0.29 | 0.37 |
| Started multiple businesses | 0.52 | 0.17 | 0.002 | 0.48 | 0.15 | 0.81 |
| *DV = Si, age and gender corrected (standardized)* |  |  |  |  |  |  |
| Never started own business | 0.02 | 0.05 | 0.701 | 0.02 | -0.07 | 0.11 |
| Started one business | 0.09 | 0.19 | 0.654 | 0.11 | -0.22 | 0.43 |
| Started multiple businesses | -0.42 | 0.17 | 0.015 | -0.40 | -0.68 | -0.11 |

Association of history of entrepreneurship, across three levels, with MMPI dimensions.

**Table S15.**
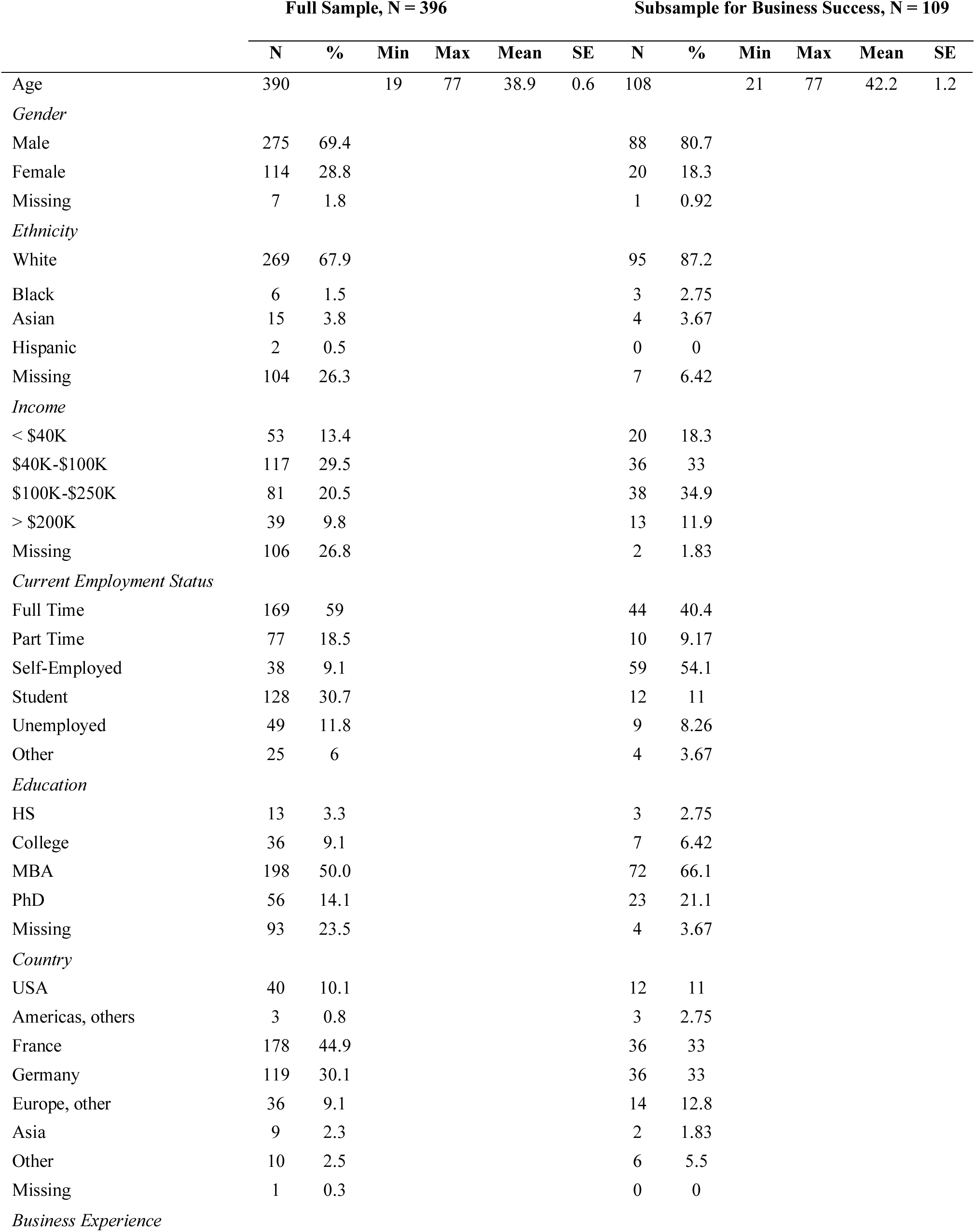
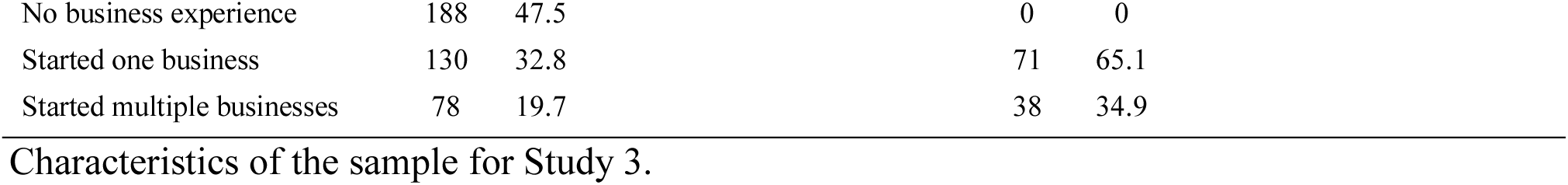

|  | Full Sample, N = 396 |  |  |  |  |  | Subsample for Business Success, N = 109 |  |  |  |  |  |
| --- | --- | --- | --- | --- | --- | --- | --- | --- | --- | --- | --- | --- |
|  | N | % | Min | Max | Mean | SE | N | % | Min | Max | Mean | SE |
| Age | 390 |  | 19 | 77 | 38.9 | 0.6 | 108 |  | 21 | 77 | 42.2 | 1.2 |
| *Gender* |  |  |  |  |  |  |  |  |  |  |  |  |
| Male | 275 | 69.4 |  |  |  |  | 88 | 80.7 |  |  |  |  |
| Female | 114 | 28.8 |  |  |  |  | 20 | 18.3 |  |  |  |  |
| Missing | 7 | 1.8 |  |  |  |  | 1 | 0.92 |  |  |  |  |
| *Ethnicity* |  |  |  |  |  |  |  |  |  |  |  |  |
| White | 269 | 67.9 |  |  |  |  | 95 | 87.2 |  |  |  |  |
| Black | 6 | 1.5 |  |  |  |  | 3 | 2.75 |  |  |  |  |
| Asian | 15 | 3.8 |  |  |  |  | 4 | 3.67 |  |  |  |  |
| Hispanic | 2 | 0.5 |  |  |  |  | 0 | 0 |  |  |  |  |
| Missing | 104 | 26.3 |  |  |  |  | 7 | 6.42 |  |  |  |  |
| *Income* |  |  |  |  |  |  |  |  |  |  |  |  |
| < \$40K | 53 | 13.4 | | | | | 20 | 18.3 | | | | |
| \$40K-\$100K | 117 | 29.5 | | | | | 36 | 33 | | | | |
| \$100K-\$250K | 81 | 20.5 | | | | | 38 | 34.9 | | | | |
| > \$200K | 39 | 9.8 | | | | | 13 | 11.9 | | | | |
| Missing | 106 | 26.8 |  |  |  |  | 2 | 1.83 |  |  |  |  |
| *Current Employment Status* |  |  |  |  |  |  |  |  |  |  |  |  |
| Full Time | 169 | 59 |  |  |  |  | 44 | 40.4 |  |  |  |  |
| Part Time | 77 | 18.5 |  |  |  |  | 10 | 9.17 |  |  |  |  |
| Self-Employed | 38 | 9.1 |  |  |  |  | 59 | 54.1 |  |  |  |  |
| Student | 128 | 30.7 |  |  |  |  | 12 | 11 |  |  |  |  |
| Unemployed | 49 | 11.8 |  |  |  |  | 9 | 8.26 |  |  |  |  |
| Other | 25 | 6 |  |  |  |  | 4 | 3.67 |  |  |  |  |
| *Education* |  |  |  |  |  |  |  |  |  |  |  |  |
| HS | 13 | 3.3 |  |  |  |  | 3 | 2.75 |  |  |  |  |
| College | 36 | 9.1 |  |  |  |  | 7 | 6.42 |  |  |  |  |
| MBA | 198 | 50.0 |  |  |  |  | 72 | 66.1 |  |  |  |  |
| PhD | 56 | 14.1 |  |  |  |  | 23 | 21.1 |  |  |  |  |
| Missing | 93 | 23.5 |  |  |  |  | 4 | 3.67 |  |  |  |  |
| *Country* |  |  |  |  |  |  |  |  |  |  |  |  |
| USA | 40 | 10.1 |  |  |  |  | 12 | 11 |  |  |  |  |
| Americas, others | 3 | 0.8 |  |  |  |  | 3 | 2.75 |  |  |  |  |
| France | 178 | 44.9 |  |  |  |  | 36 | 33 |  |  |  |  |
| Germany | 119 | 30.1 |  |  |  |  | 36 | 33 |  |  |  |  |
| Europe, other | 36 | 9.1 |  |  |  |  | 14 | 12.8 |  |  |  |  |
| Asia | 9 | 2.3 |  |  |  |  | 2 | 1.83 |  |  |  |  |
| Other | 10 | 2.5 |  |  |  |  | 6 | 5.5 |  |  |  |  |
| Missing | 1 | 0.3 |  |  |  |  | 0 | 0 |  |  |  |  |
| *Business Experience* |  |  |  |  |  |  |  |  |  |  |  |  |

|  |  |  |  |  |
| --- | --- | --- | --- | --- |
| No business experience | 188 | 47.5 | 0 | 0 |
| Started one business | 130 | 32.8 | 71 | 65.1 |
| Started multiple businesses | 78 | 19.7 | 38 | 34.9 |

Characteristics of the sample for Study 3.

**Table S16.**

|  | Descriptive Statistics |  |  |  |  | Test for normality |  | Age effects |  | Gender effects |  |
| --- | --- | --- | --- | --- | --- | --- | --- | --- | --- | --- | --- |
| | N | Mean | SE | Skew. | Kurt. | K-S | p | $\rho$ | p | Mean<sub>M</sub> - Mean<sub>F</sub> | M-W U p |
| *Hypomanic tendencies* |  |  |  |  |  |  |  |  |  |  |  |
| HPS | 396 | 20.8 | 0.4 | 0.25 | -0.17 | 0.05 | < 0.01 | -0.22 | < 0.01 | 0.55 | 0.43 |
| Social Vitality | 396 | 10.2 | 0.2 | 0.27 | -0.04 | 0.09 | < 0.01 | -0.19 | < 0.01 | 0.71 | 0.10 |
| Mood Volatility | 396 | 6.5 | 0.2 | 0.22 | -0.73 | 0.10 | < 0.01 | -0.17 | < 0.01 | -0.03 | 0.96 |
| Excitement | 396 | 2.9 | 0.1 | 0.45 | -0.53 | 0.13 | < 0.01 | -0.10 | 0.05 | -0.19 | 0.53 |
| *Entrepreneurial Outcomes Inventory* |  |  |  |  |  |  |  |  |  |  |  |
| Student Entrepreneurship | 142 | 5.0 | 0.7 | 4.60 | 27.57 | 0.29 | < 0.01 | -0.20 | 0.02 | 0.78 | 0.41 |
| Corporate Entrepreneurship | 300 | 58.2 | 12.7 | 8.63 | 87.56 | 0.39 | < 0.01 | 0.24 | < 0.01 | 32.25 | 0.01 |
| Invention Entrepreneurship | 142 | 162.1 | 148.0 | 11.86 | 141.11 | 0.50 | < 0.01 | -0.10 | 0.22 | 251.88 | <0.01 |
| *Entrepreneurial Success* |  |  |  |  |  |  |  |  |  |  |  |
| | | \$100K | \$100K | | | | | | | \$100K | |
| Annual Revenue of the most successful business | 109 | 128.1 | 92.6 | 10.06 | 103.28 | 0.45 | < 0.01 | 0.31 | < 0.01 | 0.0 | 0.99 |

Descriptive Statistics of measures of HPM and entrepreneurship in the Study 3 sample. Skew. – Skewness; Kurt. – Kurtosis; K-S - Kolmogorov-Smirnov Statistic; $\rho$ - Spearman's correlation; M-W U p - Mann-Whitney U test p-value.

**Table S17.**
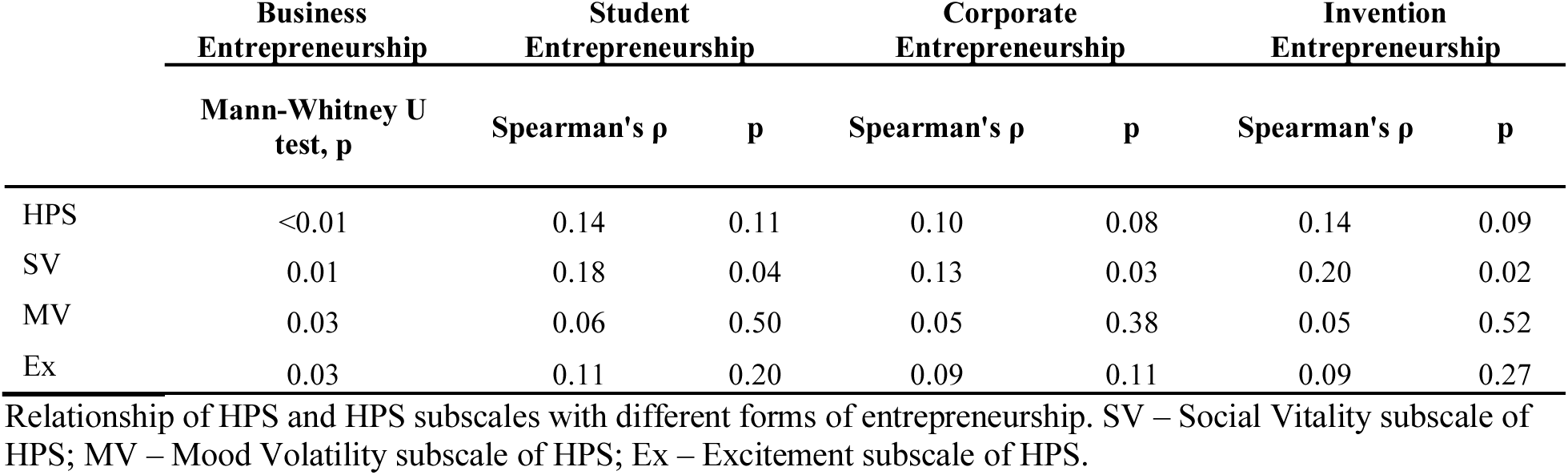

|  | **Business**<br>**Entrepreneurship** | **Student**<br>**Entrepreneurship** |  | **Corporate**<br>**Entrepreneurship** |  | **Invention**<br>**Entrepreneurship** |  |
| --- | --- | --- | --- | --- | --- | --- | --- |
|  | **Mann-Whitney U**<br>**test, p** | **Spearman's $\rho$** | **p** | **Spearman's $\rho$** | **p** | **Spearman's $\rho$** | **p** |
| HPS | <0.01 | 0.14 | 0.11 | 0.10 | 0.08 | 0.14 | 0.09 |
| SV | 0.01 | 0.18 | 0.04 | 0.13 | 0.03 | 0.20 | 0.02 |
| MV | 0.03 | 0.06 | 0.50 | 0.05 | 0.38 | 0.05 | 0.52 |
| Ex | 0.03 | 0.11 | 0.20 | 0.09 | 0.11 | 0.09 | 0.27 |

Relationship of HPS and HPS subscales with different forms of entrepreneurship. SV – Social Vitality subscale of HPS; MV – Mood Volatility subscale of HPS; Ex – Excitement subscale of HPS.

**Table S18.**
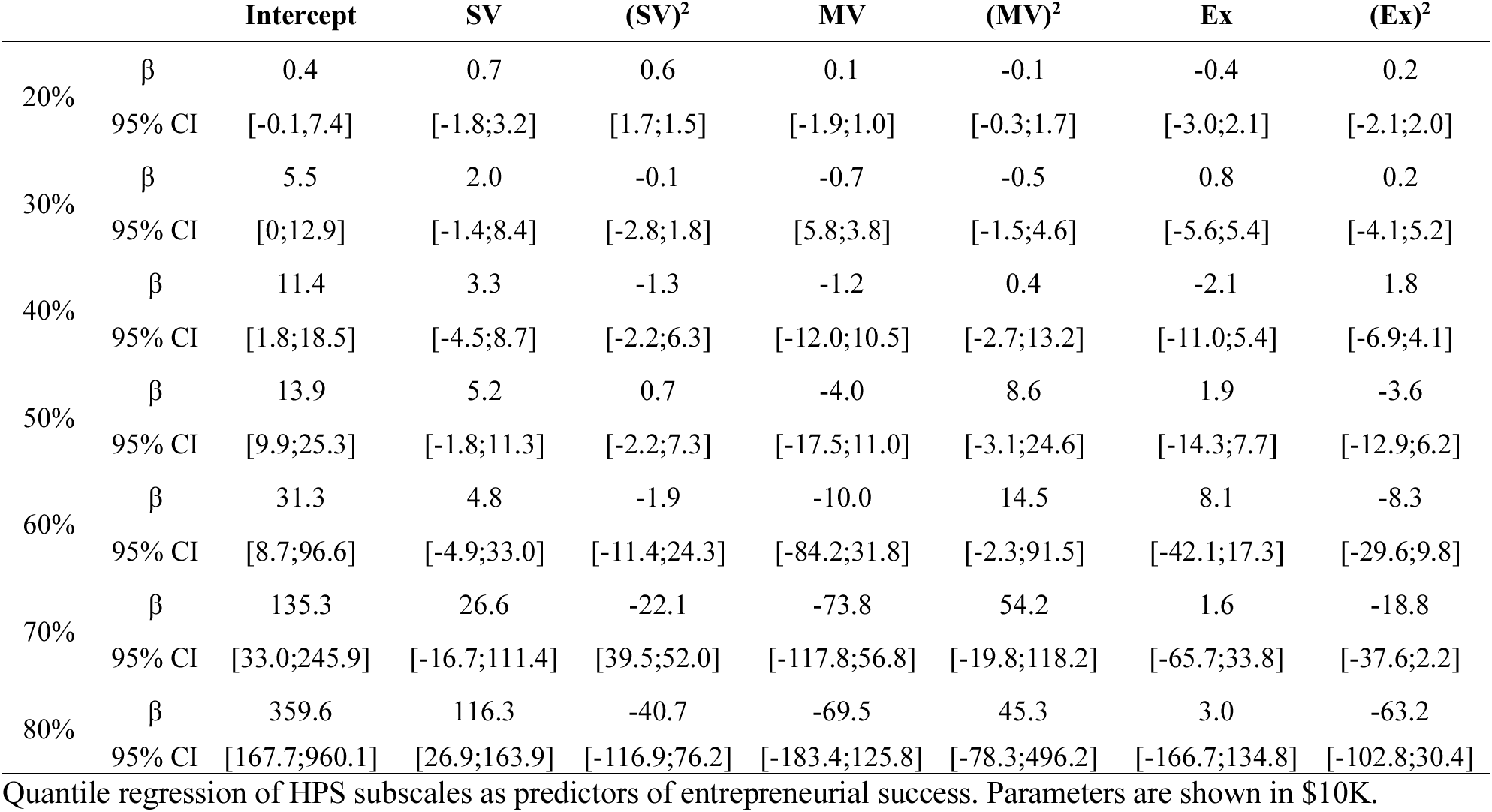

|  |  | **Intercept** | **SV** | **(SV)<sup>2</sup>** | **MV** | **(MV)<sup>2</sup>** | **Ex** | **(Ex)<sup>2</sup>** |
| --- | --- | --- | --- | --- | --- | --- | --- | --- |
| 20% | $\beta$ | 0.4 | 0.7 | 0.6 | 0.1 | -0.1 | -0.4 | 0.2 |
|  | 95% CI | [-0.1;7.4] | [-1.8;3.2] | [1.7;1.5] | [-1.9;1.0] | [-0.3;1.7] | [-3.0;2.1] | [-2.1;2.0] |
| 30% | $\beta$ | 5.5 | 2.0 | -0.1 | -0.7 | -0.5 | 0.8 | 0.2 |
|  | 95% CI | [0;12.9] | [-1.4;8.4] | [-2.8;1.8] | [5.8;3.8] | [-1.5;4.6] | [-5.6;5.4] | [-4.1;5.2] |
| 40% | $\beta$ | 11.4 | 3.3 | -1.3 | -1.2 | 0.4 | -2.1 | 1.8 |
|  | 95% CI | [1.8;18.5] | [-4.5;8.7] | [-2.2;6.3] | [-12.0;10.5] | [-2.7;13.2] | [-11.0;5.4] | [-6.9;4.1] |
| 50% | $\beta$ | 13.9 | 5.2 | 0.7 | -4.0 | 8.6 | 1.9 | -3.6 |
|  | 95% CI | [9.9;25.3] | [-1.8;11.3] | [-2.2;7.3] | [-17.5;11.0] | [-3.1;24.6] | [-14.3;7.7] | [-12.9;6.2] |
| 60% | $\beta$ | 31.3 | 4.8 | -1.9 | -10.0 | 14.5 | 8.1 | -8.3 |
|  | 95% CI | [8.7;96.6] | [-4.9;33.0] | [-11.4;24.3] | [-84.2;31.8] | [-2.3;91.5] | [-42.1;17.3] | [-29.6;9.8] |
| 70% | $\beta$ | 135.3 | 26.6 | -22.1 | -73.8 | 54.2 | 1.6 | -18.8 |
|  | 95% CI | [33.0;245.9] | [-16.7;111.4] | [39.5;52.0] | [-117.8;56.8] | [-19.8;118.2] | [-65.7;33.8] | [-37.6;2.2] |
| 80% | $\beta$ | 359.6 | 116.3 | -40.7 | -69.5 | 45.3 | 3.0 | -63.2 |
|  | 95% CI | [167.7;960.1] | [26.9;163.9] | [-116.9;76.2] | [-183.4;125.8] | [-78.3;496.2] | [-166.7;134.8] | [-102.8;30.4] |

Quantile regression of HPS subscales as predictors of entrepreneurial success. Parameters are shown in \$10K.

## Footnotes

1 Standard regression analysis yields a similar and significant relationship

